# Prolonged Water-only Fasting is a Safe and Feasible Treatment Option for Managing Stage 1 and 2 Hypertension

**DOI:** 10.1101/2024.02.04.24302309

**Authors:** Evelyn Zeiler, Sahmla Gabriel, Mackson Ncube, Natasha Thompson, Eugene Scharf, Alan C. Goldhamer, Toshia R. Myers

## Abstract

Prolonged water-only fasting appears to reduce high blood pressure but randomized controlled trials are needed. This single arm pre-post interventional trial (clinicaltrials.gov, NCT04515095) investigates the safety, feasibility, and effectiveness of prolonged water-only fasting in the treatment of stage 1 and 2 hypertension. Twenty-nine participants with stage 1 and 2 hypertension, who were pre-approved to water-only fast for ≥7 days, were enrolled from a residential fasting center. Participants received 24-hour medical supervision, and adverse events were recorded according to Common Terminology Criteria for Adverse Events (CTCAE) version 5.0. Feasibility was assessed based on retention rate, ability to complete minimal fast length, and intervention acceptability. Demographic, anthropometric, medication use, laboratory, and survey data were also collected. Twenty-seven of the 29 enrolled participants attended all study visits through the six-week follow-up visit and completed at least seven consecutive days of water-only fasting. The majority of adverse events were mild and transient and there were no higher-grade or serious adverse events. Overall, the intervention was acceptable. At the six-week follow-up visit, there were sustained reductions in median systolic/diastolic blood pressure (−19.19/-5.13 mmHg), body weight (−6.72 kg), abdominal circumference, −6.55 cm), and anti-hypertensive medication use (−93%), which also persisted at the 12-month follow-up visit. These preliminary data suggest that prolonged water-only fasting is a safe and feasible treatment option for stage 1 and 2 hypertension. The data also suggest that fasting may result in sustainable reductions in high blood pressure and anti-hypertensive medication use.

## 1 Introduction

Hypertension (HTN) affects nearly half of US American adults[1; 2; 3] and associated healthcare costs are estimated at 131 billion USD annually.[4] Uncontrolled HTN is a modifiable risk factor for developing ischemic heart disease, stroke, and other types of cardiovascular disease (CVD) as well as renal disease.[2; 3] The risk of developing CVD due to high blood pressure (BP) reportedly doubles with every 20/10 mmHg increase in systolic blood pressure (SBP)/DBP above 115/75 mmHg.[5] Current treatment guidelines recommend that all patients with high BP implement diet and lifestyle modifications and for anti-hypertensive medications to be prescribed in high-risk patients with SBP/DBP ≥130/80 mmHg (Stage 1) or ≥140/90 mmHg (Stage 2).[5] It has been reported that as many as 60% of medicated HTN patients continue to have uncontrolled high BP and that anti-hypertensive medications result in modest BP reductions and/or have low rates of adherence, potentially due to adverse side effects.[6; 7] Furthermore, anti-hypertensive medications only confer significant risk reduction in patients already at high risk for developing CVD. [8]

Although diet and lifestyle modifications may lower high BP as well as associated CVD risks and rates of polypharmacy, these interventions have proven challenging to implement and also have low adherence rates.[9; 10; 11] Preliminary research suggests that medically supervised, prolonged water-only fasting followed by an exclusively whole-plant-food diet free of added salt, oil, and sugar (SOS-Free Diet) has a very low risk of causing a severe or serious adverse event, results in immediate and sustained improvements in blood pressure and other biomarkers of CVD risk, and may improve adherence to diet and lifestyle modifications.[12; 13; 14; 15; 16] In order to continue evaluating if the intervention has potential as a HTN treatment, we conducted a single-arm pre-post interventional trial with the primary aim of assessing safety and feasibility in people with stage 1 and 2 hypertension by systematically collecting and reporting adverse events and treatment deviations as well as study retention rate, ability to complete minimal fast length, and self-reported intervention acceptability. Additionally, we measured the immediate, sustained, and long-term effects of the full intervention on BP and other cardiometabolic risk factors.

## 2 Methods

### Ethical Approval

This study was approved by the TrueNorth Health Foundation Institutional Review Board (TNHF2020-1HTN and TNHF2021-1HTNFU) and registered at clinicaltrials.gov (NCT04515095). The research was conducted in accordance with the approved protocol and complied with the standards of the Declaration of Helsinki. All participants provided written informed consent before data collection began.

### Participant Enrollment and Study Protocol

This study was a single-arm pre-post intervention trial that recruited patients with stage 1 and 2 hypertension who were undergoing an elective, medically supervised, water-only fast of at least seven days at a residential fasting center between August 2020 and September 2021. Thirty participants were enrolled, of which one was ineligible due to preexisting hyponatremia that contraindicates fasting (**Figure 1**, **ST1**). Consenting participants of any sex, aged 30-75 years, with uncontrolled (SBP ≥130 mmHg and/or DBP ≥80 mmHg) or medication-controlled hypertension, fasting glucose <126 mg/dL and/or hemoglobin A1c <7%, and prior approval by a non-research physician to water-only fast for at least seven consecutive days followed by a refeeding period of at least half the fasting length, were eligible for inclusion. Exclusion criteria included SBP >180 and/or DBP >120 mmHg at time of enrollment, active malignancy, active kidney disease, active inflammatory disorder (including classic autoimmune connective tissue disorders, multiple sclerosis, inflammatory bowel disorders), stroke or heart attack within the last 90 days, and the inability to discontinue medications and/or supplements. Study dates overlapped with NCT04514146, conducted at the same fasting center, and 18/29 participants were co-enrolled in that study.[15]

**Figure 1.** Enrollment and participation flow diagram. See “Participant Enrollment and Study Protocol” in the “Materials and Methods” section and “Participant Characteristics” in the “Results” section for details on eligibility and data collection. BL, baseline; EOF, end-of-fast; EOR, end-of-refeed; 6wFU, six-week follow-up; 12mFU, 12-month follow-up.

**Figure 2.** SBP (A, C, E) and DBP (B, D, F) by visit of all (A, B), unmedicated (C, D) and medicated (E, F) participants at BL. Circles and squares in panels **C**, **D**, **E**, **F** represent unmedicated and medicated SBP/DBP, respectively. Participants who attended the 12mFU visit (n=17) are depicted as closed (black) symbols. Horizontal dotted lines represent severity of HTN: SBP 120-129 mmHg, elevated blood pressure; SBP/DBP ≥ 130/80 mmHg, Stage 1 HTN; SBP/DBP ≥ 140/90 mmHg, Stage 2 HTN. 3/29 and 12/29 participants did not provide data at 6wFU and 12mFU visits, respectively. SBP, systolic blood pressure; DBP, diastolic blood pressure; mmHg, millimeter mercury; BL, baseline; EOF, end-of-fast; EOR, end-of-refeed; 6wFU, six-week follow-up; 12mFU, 12-month follow-up; HTN, hypertension.

Demographic information, medical diagnoses, and current use of anti-hypertensive medications were collected at BL, 6wFU, and 12mFU visits (see below). During the treatment period, onsite study visits occurred daily and at baseline (BL), end of fast (EOF), and end of refeed (EOR). Two additional study visits occurred either onsite or remotely: an expected visit that occurred six weeks after departure from the fasting center (6wFU) and an unexpected visit that occurred 12 months after the 6wFU visit (12mFU). During daily visits, body weight (BW), BP, peripheral oxygen saturation (SpO_2_), body temperature (BT), and pulse were measured and adverse events (AEs) were recorded. Additionally, BW, BP, abdominal circumference (AC), and blood and urine samples were collected at BL, EOF, EOR, 6wFU, and 12mFU visits (see below). Web-based versions of three questionnaires were administered: the SOS-Free Diet Screener[15] at BL, 6wFU, and 12mFU visits; the Treatment Adherence/Acceptability Scale (TAAS)[17] questionnaire at EOF and 6wFU visits; and the Food Acceptability Questionnaire (FAQ)[18] at BL, EOR, 6wFU, and 12mFU visits. Details about the questionnaires can be found in the supplemental material. Demographic information, medical diagnoses, and current use of anti-hypertensive medications were collected at BL, 6wFU, and 12mFU visits (see Supplemental Material). Data collection and survey distribution were performed using the web-based software Research Electronic Data Capture (REDCap).[19] Missing data are described in **ST1** and in the legend of respective tables.

### Medically Supervised, Water-only Fasting Protocol

The prefeeding, water-only fasting, and refeeding protocol took place at a residential fasting center. The protocol was previously described in detail [14]. Participants were approved to water-only fast by medical doctors not affiliated with this study after a thorough examination, which included detailed patient history, comprehensive physical exam, basic neurological and psychological status, complete blood count (CBC), comprehensive metabolic panel (CMP), urinalysis, and additional tests as clinically indicated. Medication use while water-only fasting is contraindicated and ability to discontinue medication use safely was a requirement for enrollment in this study. Participants received 24-hour medical supervision during the entire treatment (i.e., prefeeding, water-only fasting, and refeeding). Vital signs were examined by medical personnel twice per day and labs were ordered once per week or as requested by the attending physician.

#### Prefeeding

A prefeeding period began at least two days prior to the water-only fast, during which participants eliminated all recreational drugs (e.g., coffee, alcohol, nicotine, etc.) and specific foods (i.e., grains, legumes, dairy, meat, seafood, added sugar, oils and salt, and all processed foods) and ate only the provided raw/steamed fruits and vegetables. During this time, participants were weaned off anti-hypertensive and other medications as determined appropriate by the attending physician based on clinical protocol and according to the individual participant’s medical needs.

#### Fasting

During the water-only fast, participants drank a minimum of 1.2 L/day of steam-distilled water. If medically indicated (e.g., due to blood electrolyte imbalance, hypoglycemia, or gastroesophageal reflux disease) or desired by the participant, vegetable broth (VB; 80 kcal per day) was consumed. VB has such an insignificant amount of calories that it does not prevent ketosis or reverse ketosis once it has been initiated and is assumed equivalent to water. The fast continued for the predetermined amount of time or modified according to the individual patient’s needs. In cases of more severe adverse events or discomfort, the fast was temporarily or indefinitely broken with fruit and vegetable juice (500-600 kcal per day) or potato/zucchini blend (900 kcal per day).

#### Refeeding

The refeeding diet was administered in five phases: 1) fruit and vegetable juice; 2) raw fruits/vegetables; 3) raw/steamed vegetables and fruits; 4) raw/steamed fruits and vegetables, grains, and nuts or avocado; 5) unrestricted whole-plant foods free of added salt, oil, and sugar. Each of the five refeeding phases lasted one day for every 7-10 days of water-only fasting, for a period of time lasting at least one half of the total fast length (e.g., 10 days of fasting would correspond to five days of refeeding with one day on each of the five phases). If medically indicated (e.g., due to food intolerance), the refeeding protocol was modified.[14] Before leaving the center, participants received basic nutrition education and were instructed to continue the exclusively whole-plant-food, SOS-free diet for at least the next six weeks. There were no further instructions after the 6wFU visit.

### Adverse Events Recording & Analysis

While prefeeding, fasting and refeeding, AEs were identified in daily interviews conducted by trained clinical research personnel, daily physical (i.e., daily vitals) examinations, weekly blood analysis (i.e., CBC and CMP), and weekly urinalysis. AEs were recorded by date and according to Common Terminology Criteria for Adverse Events (CTCAE)[20] based on symptom categorization and severity (grade 1, mild; grade 2, moderate; grade 3, severe; grade 4, life-threatening; grade 5, death). The CTCAE describes AE terms according to the Medical Dictionary for Regulatory Activities (MedDRA), as lowest-level terms (LLT) that are grouped into a system organ class (SOC). For terms not specifically listed in the CTCAE and listed under the MedDRA SOC as ‘other’, new terms were added and graded according to the CTCAE guideline that described the AE as best as possible. All AEs were reported regardless of attribution. Pre-existing conditions were considered an AE if reoccurring or increasing in severity at any time during the intervention. The same AE may have occurred multiple times for the same participant and was counted as a new event if it decreased or increased in grade or had previously resolved but subsequently returned. AE outcomes (i.e., resolved, persisted, unknown) were determined as follows: the outcome was considered “resolved” if the symptom cleared completely or the severity of the event decreased (e.g., G2 decreased to G1). The outcome was considered “persisted” if the AE did not resolve at EOR or 6wFU visit. The outcome was considered “unknown” if the interview or specific analysis was not repeated (only AEs determined by CMP and CBC were monitored at 6wFU) or if the participant failed to report back at the 6wFU visit. The total number of AEs for all participants during the entire intervention was counted and presented based on SOC, LLT, grade, and outcome. Additionally, the top 10% of total AEs experienced by participants were presented based on LLT, grade, and treatment phase (i.e., prefeeding, fasting, and refeeding). AEs were further categorized as the highest-grade adverse event (HGAE) during the entirety of each participant’s intervention.

### Demographics, Medical Diagnoses, Medication Use, and Anthropometric Measurements

Demographic information (age, sex), current and past medical diagnoses, and current medication use was collected by interview at BL and self-reported at 6wFU and 12mFU visits using an online survey. Height (cm) was measured once at BL with a digital wall mounted stadiometer (DS5100, Doran Scales Inc., St. Charles, IL, USA) and used to calculate body mass index (BMI) at each time point. Participants were instructed to wear only one layer of clothing and remove shoes and any pocket items for BW measurements. BW (kg) was measured using a digital body scale (BWB 800A Class III, Tanita Corporation of American Inc., Arlington Heights, IL, USA) while onsite and with the provided digital floor scale (WW26, Conair LLC, Stamford, CT, USA) while offsite. AC (cm) was measured horizontally on a bare abdomen at the minimal waistline and parallel to the floor with a tension-sensitive, non-elastic tape (Gullick II, Model 67019, Country Technology Inc., Gay Mills, WI, USA) while onsite or with the provided retractable tape measure while offsite. The measurement was read in the midaxillary line and at the end of a normal expiration. BP was measured while the participant was resting in a seated position and their left arm elevated to heart level. A digital blood pressure device (Welch Allyn-Connex ProBP 3400, Hill-Rom Holding Inc. Chicago, IL, USA) was used while onsite and a blood pressure monitor (BP3GX1, Microlife USA Inc, Clearwater, FL, USA) for the offsite visits. Participants received thorough education as well as written instructions in order to provide self-reported measurements at 6wFU and 12mFU visits.

### Biospecimen Collection

For onsite visits, serum and plasma blood samples as well as 24-hour urine were collected and sent to LabCorp (LabCorp, Burlington, NC, USA). For offsite visits, the samples were collected directly at a LabCorp local to the participant. All blood samples were drawn by a certified phlebotomist. Blood samples were drawn while onsite by a certified phlebotomist at BL, EOF, and EOR and at a local LabCorp for the off-site 6wFU and 12mFU visits. Blood collection took place in the morning, before the consumption of caloric food or liquid but with the instruction to consume 1-2 cups of water prior to arrival. While in a seated position, blood was collected into one red top (Red top, 16 × 100, 10 mL, silica, BD, Mississauga, ON, Canada) and one lavender top vacutainer tube (Lavender top, 13×75, 4.0 ml, K2EDTA, BD, Mississauga, ON, Canada). The lavender top vacutainer was placed into an ice/water bath immediately after collection and stored at 4^◦^C until it was sent to a commercial laboratory (LabCorp, Burlington, NC, USA) for complete blood count (CBC) analysis. LabCorp reports that CBC analysis was completed using an Auto cell counter with mixed technologies from Sysmex (Sysmex Asia Pacific Pte Ltd, Asia Green, Singapore). The red top vacutainer tube was incubated at room temperature for 30-50 min and centrifuged at 1500g for 10 minutes at 4°C (Model: Thermo Scientific Sorvall Legend X1R, Rotor: TX-400 Rotor Cross, Thermo Fisher Scientific Inc., Waltham, MA, USA). The serum was separated and stored at 4^◦^C until it was sent to LabCorp for comprehensive metabolic panel (CMP) analysis, lipid panel (total cholesterol, HDL-, LDL-, VLDL-cholesterol, triglycerides), C-reactive protein (CRP), gamma-glutamyl transferase (GGT), and insulin. LabCorp reports that CMP was measured using Ion Selective Electrode (ISE), colorimetric, enzymatic, and kinetic methods from Roche Diagnostics (Roche Diagnostics, Indianapolis, IN, USA), lipid panel by colorimetric and enzymatic methods, insulin by electrochemiluminescence immunoassays (ECLIA), CRP by high sensitivity immunochemiluminometric assays (ICMA), and GGT by kinetic methods from Roche Diagnostics (Roche Diagnostics, Indianapolis, IN, USA).

Participants collected 24-hour urine samples at BL, EOF, EOR, 6wFU, and 12mFU visits. The collection started after the first void was discarded and subsequent voids were collected into one or two additive-free urine containers (LabCorp) and stored at 4^◦^C for the next 24 hours. After completion, 24-hour urine samples were combined and/or inverted to ensure homogeneity and the total volume was assessed. 50 ml was transferred into a cup and a dipstick urinalysis (Siemens Multistix 10SG [Siemens Medical Solutions USA, Inc., Malvern, PA, USA] or Mission Urinalysis Strips [ACON Laboratories, Inc., San Diego, CA, USA]) was performed at BL, EOF, and EOR to measure leukocytes, nitrite, urobilinogen, protein, pH, blood, specific gravity, ketone, bilirubin, and glucose.

BMI (kg/m^2^) was calculated using the formula[21]: weight (kg) ÷ height^2^ (m^2^); the homeostatic model assessment of insulin resistance (HOMA-IR) was calculated using the formula[22]: insulin (µU/L) × glucose (nmol/L)/22.5; and fatty liver index (FLI) was calculated using the formula[23]:

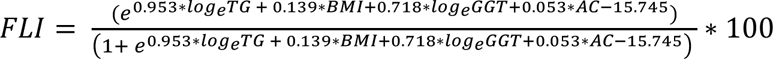

### Daily Vitals

Daily vitals were collected onsite by trained clinical research staff every morning from BL through EOR. Participants were asked to rest in a seated position with their arms elevated to heart level on a table or pillow for five minutes prior to having their BP (Welch Allyn-Connex ProBP 3400, Hill-Rom Holding Inc. Chicago, IL, USA), BT (Welch Allyn Sure Temp plus 690, Hill-Rom Holding Inc. Chicago, IL, USA), peripheral oxygen saturation (SpO2, Zacurate 500DL, Zacurate, Stafford, TX, USA), and pulse rate and rhythm (manually for 15 seconds or 1 min) measured. BW was self-reported using a floor scale (WW26, Conair LLC, Stamford, CT, USA). Participants were also asked about hours of sleep, energy level, urination characteristics (dysuria, difficulty, changes), nausea, presyncope, quantity of water intake, bowel movements (frequency, characteristics), and any other symptoms for the previous 24 hours as well as about any unresolved complaints.

### Questionnaires

#### SOS-Free Dietary Screener

Adherence to the recommended SOS-Free Diet was assessed with a 27-question screener as previously described.[15] The SOS-Free Dietary Screener was administered online at BL, 6wFU, and 12mFU visits. A non-adherence score was calculated based on consumption of inclusionary and exclusionary foods over the previous 30 days. The minimum non-adherence score is zero (i.e., 100% adherent) and the maximum non-adherence score is 82.5 (i.e., 0% adherent).

#### Treatment Acceptability Adherence Scale

Acceptability of the treatment was assessed using the Treatment Acceptability/Adherence Scale (TAAS).[17] The TAAS was administered online at EOF and 6wFU visits. The TAAS contains 10 questions with response options ranging from 1 (disagree strongly) to 4 (neither agree nor disagree) to 7 (agree strongly). Negatively worded questions (#3-5, #7-8, #10) were reverse scored and total scores were obtained by summing all answers. Scoring ranged from 70 to 10 with higher scores indicating greater treatment acceptability. Individual questions are shown **SF1**.

#### Food Acceptability Questionnaire

Acceptability of foods permitted on the SOS-Free Diet was assessed using the Food Acceptability Questionnaire.[18] The FAQ was administered online at BL, EOR, 6wFU, and 12mFU visits. The questionnaire contains 10 questions with response options ranging from 1 (not at all) to 7 (extremely). Negatively worded questions (#4 and #8) were reverse scored and total scores were obtained by summing all answers. Scoring ranged from 70 to 10 with higher scores indicating greater acceptability of permissible foods. Individual questions are shown in **SF2**.

### Statistical Analysis

This single-arm pre-post interventional trial evaluates changes in clinical parameters across visits. The absence of a control group inhibits the ability to discern if observed changes result from the intervention, natural progression, external influences, Hawthorne effects, or other factors. Data cleaning and statistical analysis were conducted using R (R Core Team, 2022), with descriptive statistics reported as medians and interquartile ranges (IQR).[24] Bivariate relationships between clinical parameters were visualized through scatter plots. The statistical analysis aimed to estimate within-group changes between visits (e.g., BL, EOF, EOR, 6wFU, and 12mFU) and identify significant changes. The analysis employed random intercept generalized linear mixed-effects models, with clinical parameters as dependent variables and participant ID as the grouping variable. Age and sex served as fixed effect control variables in all models; baseline hypertension status was also included for binary outcome models.

Models for continuous outcomes were fitted using lmerTest version 3.1.3,[25] while binary and count outcomes used lme4 version 1.1.32.[26] Binary data were modeled using a binomial family parameter with a logit link, and count data used a Poisson family parameter with a log link. Regression coefficients for all clinical parameters, along with 95% confidence intervals (CI), were derived from broom.mixed version 0.2.9.4.[27] Significance for continuous data was determined when CIs excluded zero, and for binary and count data, when exponentiated CIs excluded one. Missing data were addressed through complete case analysis where case is defined as a visit within participant. Model diagnostics employed graphical methods via the modelDiagnostics function from multilevelTools version 0.0.1.[28] Where diagnostics indicated model assumption violations, results were compared between lmerTest and the Robust Scoring Equations estimator (RSE) from robustlmm version 3.2.0[29; 30] with the Satterthwaite degrees of freedom approximation from sjPlot version 2.8.14.[31] Discrepancies in effect magnitude, direction, or significance led to RSE results being prioritized. The RSE method was not available for binary or count outcomes.

After data collection was complete, a post-hoc simulation-based power analysis[32] was conducted. This analysis used hypothetical population effect sizes and standard deviations[33] derived from the SBP and BMI models in Table 4 of Gabriel et al.[15] The goal was to estimate the sample size required to achieve 80% statistical power at an alpha level of 0.05 for parameter estimates assessing within-group changes from BL to 6wFU visits. For SBP, the power analysis indicated that 80% power could be attained with 32 participants. This calculation was based on data simulated from a model with population effect sizes set as follows: −11.49 mmHg for EOF – BL, −14.58 mmHg for EOR – BL, −7.68 mmHg for 6wFU – BL, and −1.92 mmHg for 12mFU – BL. Since, Gabriel et al.[15] did not include a 12mFU visit, the 12mFU effect size of −1.92 mmHg represents a 25% reduction from the 6wFU effect. Additional model parameters included 0.21 mmHg per year for age and 0.73 mmHg for sex. The population standard deviations were set at 10.58 mmHg for the group level and 10.55 mmHg for the residual standard deviation. Adjusting the simulation to increase the magnitude of effect sizes associated with the visits by 10%, while maintaining other effect sizes and standard deviations as in the initial simulation, resulted in achieving 80% power with a reduced sample size of 27 participants. For the exploratory secondary endpoint BMI, the analysis demonstrated that 80% statistical power could be reached with 27 participants. This was based on a model simulation with population effect sizes as follows: −1.92 kg/m² for EOF – BL, −1.61 kg/m² for EOR – BL, −1.62 kg/m² for 6wFU – BL, and −0.24 kg/m² for 12mFU – BL. The model also included age and sex effect sizes of −0.01 kg/m² per year and −1.10 kg/m², respectively. The population standard deviations were set at 7.07 kg/m² for the group level and 1.08 kg/m² for the residual standard deviation. These parameters represented a 60% reduction in the magnitude of effect sizes associated with the visits and a 150% increase in the standard deviations, in comparison to the model that produced the BMI results in Table 4 of Gabriel et al.[15]

The rates of change in clinical parameters were estimated using linear mixed-effect models[34] implemented with Linear and Nonlinear Mixed Effects Models (nlme) version 3.1.160.[35] In these models, the clinical parameter was designated as the dependent variable, the day of the study phase as the fixed effect, and participant ID as the random intercept. Additionally, the fully specified model included the day of the study phase as the random slope. This fully specified model was preferred when both the Akaike Information Criterion (AIC) and Bayesian Information Criterion (BIC) were lower compared to the parsimonious model, which did not include the random slope. The parameter estimate for the day of study phase fixed effect was interpreted as the rate of change for each clinical parameter. Separate models were fitted for distinct study phases: prefeeding, fasting, and refeeding. For the body weight clinical parameter, two periods of fasting were analyzed: early and late fasting. Estimates for body weight during these fasting periods were derived from a linear mixed effects model that included day of study phase, period of study phase, and their interaction as fixed effects. Participant ID was set as the random intercept and day of study phase as the random slope in the model. The early fasting period included days 1 to 5 and the late fasting period included the subsequent days. To assess homogeneity of variance and normality of residuals, residual plots and quantile-quantile plots were utilized, respectively. When violations of these assumptions were notable, RSEs[29] were used with the Satterthwaite degrees of freedom approximation.[31]

## 3 Results

### Participant Characteristics

Of the 29 eligible participants, there were 19 females and 10 males with a combined median (IQR) age of 62 (58, 67) years (**Table 1**). At the BL visit, all participants had a diagnosis of stage 1 or 2 hypertension (HTN) and 15 (52%) were taking antihypertensive medications. Five (17%) participants had medication-controlled SBP/DBP of <130/<80 mmHg, seven (24%) had a SBP and/or DBP between 130-139/80-89 mmHg (stage 1), and 17 (59%) had a SBP and/or DBP ≥140/≥90 mmHg (stage 2; **Table 1**). The most common preexisting comorbidity was mixed and unspecified hyperlipidemia (n=12; 45%) (**ST2**). At the BL visit, four (14%) participants were normal weight, 10 (34%) were overweight, and 15 (52%) were obese (**Table 1**). At the 12mFU visit there were 17 participants, of which, 12 were female and five were male with a median (IQR) age of 63 (60, 69) years.

**Table 1.** Select Baseline Characteristics.

|  |  |
| --- | --- |
| Total Participants, N | 29 |
| Total Medicated<sup>^</sup> Participants, n (%) | 15 (52) |
| Female, n (%) | 19 (66) |
| Median (IQR) Age, y | 62 (58, 67) |
| Medicated<sup>^</sup> SBP/DBP = <130/80 mmHg, n (%) | 5 (17) |
| SBP/DBP<sup>*</sup> = 130-139/80-89 mmHg, n (%) | 7 (24) |
| SBP/DBP<sup>+</sup> = ≥140/90, n (%) | 17 (59) |
| Normal Weight BMI, n (%) | 4 (14) |
| Overweight BMI, n (%) | 10 (34) |
| Obese BMI, n (%) | 15 (52) |

N, the total number of participants; n, a subset of 29 participants; %, percentage; IQR, interquartile range; y, years; SBP, systolic blood pressure; DBP, diastolic blood pressure: Stage 1 HTN = 130-139/80-89 mmHg, Stage 2 HTN = ≥ 140/90 mmHg; BMI, body mass index: normal weight, 18.5-<25 kg/m<sup>2</sup>; overweight, 25-<29 kg/m<sup>2</sup>; obese ≥ 30 kg/m<sup>2</sup>. <sup>^</sup>Medicated with anti-hypertensive medications. <sup>\*</sup>2/7 participants were taking anti-hypertensive medications. <sup>+</sup>8/17 participants using anti-hypertensive medications.

### Treatment & Visit Characteristics

Participants stayed onsite at a residential fasting center in order to receive medical supervision while prefeeding, fasting, and refeeding for a median (range) of 2 (1-4), 11 (7-40), and 5 (3-17) days, respectively (**ST3)**. Participants were instructed to continue eating an SOS-free diet while offsite between EOR and 6wFU visits and did not receive further dietary instruction thereafter. The 6wFU and 12mFU visits occurred at a median (range) of 45 (40-58) days after the EOR visit and 364 (358, 383) days after the 6wFU visit, respectively (**ST3**). All participants completed BL, EOF, and EOR visits (**Figure 1**, **ST1**), of which 2/29 participants had partially incomplete data at EOF. Twenty-seven of the 29 participants attended the 6wFU visit, of which 2/27 had partially incomplete data and 17/27 consented to attend the 12mFU visit (**ST1**). Of the 17 returning participants, five voluntarily visited TNHC for additional interventions between 6wFU and 12mFU visits. Of these, two participants visited TNHC one time and three participants visited two times, with a median (range) fasting and refeeding length of 10 (8, 13) and 6 (4, 9) days, respectively. (ST**1**, **ST3**).

### Water-only Fasting and Refeeding Treatment Acceptability & SOS-Free Dietary Adherence

Participant acceptability of the entire fasting and refeeding intervention (BL to EOR) was assessed using the Treatment Acceptability/Adherence Screener (TAAS) with scores ranging from 10 (no acceptability/adherence) to 70 (full acceptability/adherence). Responses to individual TAAS questions are shown in **SF1**. Median (IQR) scores were 67 (63, 70) and 66 (60, 69) at EOF and 6wFU visits, respectively.

Participant acceptability of the foods permitted on the SOS-Free Diet was assessed using the Food Acceptability Questionnaire (FAQ) with scores ranging from 10 (no acceptability) to 70 (full acceptability). Responses to individual FAQ questions are shown in **SF2**. Median (IQR) scores were 47 (44, 52), 49 (45, 58), 48 (42, 55), and 45 (32, 70) at BL, EOR, 6wFU, and 12mFU visits, respectively.

Participant adherence to the SOS-Free Diet during the six-week follow-up period was assessed using a previously described dietary screener with scores ranging from zero (100% adherent) to 82 (0% adherent).[15] Responses to individual questions are in **ST4**. The mean (SD) score was 12 (10), 6 (3), and 6 (4) at BL, 6wFU, and 12mFU, respectively.

Overall, these results suggest that participants found fasting and refeeding at the residential fasting center to be highly acceptable. The recommended diet was found to be moderately acceptable and participants reported imperfect adherence that improved only slightly at 6wFU and 12mFU visits compared to the period before the BL visit. Participants primarily increased fruit and vegetable consumption and decreased animal products, added salt, and oil.

### Adverse Events Identified During Prefeeding, Fasting, and Refeeding

Daily interviews and physical examinations as well as weekly hematology, serology, urinalyses, and additional testing as indicated were used to identify AEs through the EOR visit. AEs were classified into categories and grades using CTCAE v.5.0.[20] We identified 453 AEs, of which 381 (84%) were mild Grade 1 (G1), 64 (14%) were moderate Grade 2 (G2), and 8 (2%) were severe Grade 3 (G3) events (**Table 2**). There were no Grade 4 (life-threatening), Grade 5 (death), or serious AEs. There were 11 (2%), 350 (77%), and 92 (20%) AEs during prefeeding, fasting, and refeeding, respectively (**Table 2**). G1, G2, and G3 were the highest grade AE (HGAE) experienced by 7% (2/29), 69% (20/29), and 24% (7/29) of participants, respectively.

**Table 2.** Total Adverse Events by Grade & Treatment Phase.

| AE Grade | N (%) |  |  |  |
| --- | --- | --- | --- | --- |
|  | Prefeeding | Fasting | Refeeding | All Phases |
| 1 | 9 | 289 | 83 | 381 (84) |
| 2 | 2 | 54 | 8 | 64 (14) |
| 3 | 0 | 7 | 1 | 8 (2) |
| 4 | 0 | 0 | 0 | 0 |
| 5 | 0 | 0 | 0 | 0 |
| Total | 11 (2) | 350 (77) | 92 (20) | 453 (100) |

N (%), number and percentage of total adverse events; AE, adverse event; Grade 1, mild; Grade 2, moderate; Grade 3, severe; Grade 4, life-threatening; Grade 5, death. There were no Grade 4 or 5 events.

**Supplemental Table 5** shows AEs experienced by >10% of participants (N=29) by treatment stage. Overall, the top five events were mild-to-moderate fatigue, mild decrease in blood bicarbonate, mild decrease in BUN/Creatinine, mild-to-moderate high blood pressure, and mild-to-moderate nausea (**ST5**, **ST6**). The three most frequently occurring events during fasting were fatigue, hypertension, and decreased blood bicarbonate. There were 15 AE classifications that only occurred during fasting (**ST5)** including presyncope and changes in blood chemistry and/or blood count identified by CMP and CBC analysis (**ST7 - ST10**). During refeeding, fatigue was the most common event and diarrhea occurred only during refeeding (**ST5**). As expected, 4/5 hyperglycemia events occurred during refeeding. The other hyperglycemic event was identified at the EOF visit and may be due to an unreported protocol deviation.

Total AEs, associated system organ class, and event outcome are shown in **ST6**. The most frequently occurring G1 events were fatigue (n=60/381), dizziness (n=23/381), decreased blood bicarbonate (n=22/381), decreased BUN/creatinine ratio (n=18/381), nausea (n=13/381), and decreased chloride (n=13/381). Of the 381 G1 events, 33 were unresolved at EOR or 6wFU visits, and the outcome of 19/381 G1 events were unknown due to either unrepeated tests (9/19) or participant drop out (10/19).).. The most frequently occurring G2 events were fatigue (n=19/64), hypertension (16/64), and presyncope (n=10/64). All of the G2 events resolved while onsite (n=63/64) or before the 6wFU visit (n=1/64). Of the eight G3 events, seven were hypertensive events and one was a low neutrophil count. All G3 hypertensive events resolved before the EOR visit, but it is unknown if the low neutrophil count resolved due to participant drop out at the 6wFU visit.

### Deviations in Water-only Fasting

Twenty-seven of 29 participants (93%) completed at least seven consecutive days of fasting, of these 17/27 (63%) fasted solely on water, 9/27 fasted on water and vegetable broth (VB), and 1/27 (4%) fasted only on VB **(ST3)**. Fasting on VB provides only 80 kcal/day and does not affect ketosis. Therefore, it is considered similar to water-only fasting. Of the 10 deviations with VB, at least six were due to a treatment emergent AE, which included G2 presyncope (n=3), G1 gastroesophageal reflux (n=1), G1 hypokalemia (n=1), and G1 dry mouth (n=1). The one participant who fasted entirely on VB did so because of preexisting cardiac arrhythmia. Of the two participants who did not complete at least seven consecutive days of fasting, one temporarily interrupted the fast with potato/zucchini blend due to a G2 gastroesophageal reflux event and one modified the fast with vegetable and fruit juice due to G2 hypoglycemia **(ST3)**. Additionally, two of the 27 who completed at least seven consecutive fasting days had their fast prematurely terminated after day seven due to an AE, which included one participant with a G2 nausea event and one with concurrent G1 hypokalemia, and G1 electrocardiogram QT corrected prolonged events.

### Additional Hematology, Serology, and Urinalysis for AE recording

In addition to what is normally collected according to clinical protocol, CBC and CMP were performed at BL, EOF, and EOR visits. CBC and CMP were also analyzed at 6wFU and 12mFU visits. AEs resulting from abnormal CBC, CMP, and urinalysis values for individual participants are described above and in **ST5**, **ST6**. Throughout the study, median values for CBC variables remained within normal range (**ST7**, **ST8**). Median (IQR) CMP values reached abnormal values and included low BUN [6 (4, 8) mg/dL] for participants >60 years old at the EOR visit, low BUN/creatinine ratio at EOF [8 (7, 11) mg/dL] and EOR [7 (6, 9) mg/dL] visits for the entire population, and decreased carbon dioxide [17 (15, 20) mmol/L] at the EOF visit (**ST9**, **ST10**). Median values increased to the normal range for carbon dioxide at the EOR visit and for BUN and BUN/creatinine ratio at the 6wFU visit. Significant estimated changes for some CMP variables may be clinically meaningful even though the median value remained within normal limits. For example, baseline sodium significantly decreased by an estimated (95% CI) −2.79 (−4.06, −1.53) mmol/L by the end of fasting and there were four transient grade 1 hyponatremia events during fasting, but the end-of-fast median (IQR) sodium level of 141 (139, 143) mmol/L remained within normal limits (134-144 mmol/L). Twenty-four-hour dipstick urinalysis was also assessed weekly by the attending physician to monitor fasting as well as at BL, EOF, and EOR visits. As expected, ketones increased and pH decreased during fasting but reverted to baseline upon refeeding. Values for other variables were unremarkable (**ST11**).

### Changes in BW, BMI, AC, and Vital Signs

There was an average BW loss of −0.54 kg/day while fasting and an average gain of 0.23 kg/day while refeeding (**ST12**, **SF3A**). Accordingly, baseline median (IQR) BW reduced from 86.6 (80.1, 97.6) kg to 79.1 (72.4, 87.8) kg and 81.4 (73.8, 88.7) kg at EOF and EOR visits, respectively (**Table 3**, **Figure 4A**). The estimated loss (95% CI) between BL and 6wFU visits was −6.72 (−7.87, −5.58) kg, which was sustained at the 12mFU visit (**Table 3**, **ST13**). Median BMI significantly decreased from 31.0 (26.5, 33.2) kg^2^/m^2^ at the BL visit to 28.4 (23.5, 31.1) kg^2^/m^2^ at the 12mFU visit (**Table 3, ST13, Figure 4B**). The number (%) of participants that were normal weight, overweight, and obese was 4 (14), 10 (34), and 15 (52) at the BL visit and 5 (29), 7 (41), 5 (29) at the 12mFU visit, respectively (**ST14**). In females, baseline median (IQR) AC reduced from 100.5 (91.8, 108.7) to 91.5 (86.7, 101.0) at the EOF visit and to 90.6 (85.0, 95.0) cm at the 12mFU visit (**Figure 4C**, **Table 3, ST13**). In males, baseline median (IQR) AC reduced from 100.3 (95.7, 107.3) cm to 90.8 (85.8, 100.0) at the EOF visit and to 91.0 (81.3, 95.0) cm at the 12mFU visit. Overall, this corresponded to an estimated AC loss (95% CI) of −7.92 (−9.58, −6.26) cm between BL and 12mFU visits (**ST13**). Pulse, SpO2, and BT were measured daily while participants were onsite (**ST12, ST15**). There was a significant increase in median pulse from BL to EOF visits corresponding to an estimated rate of change of 0.55 beats/min per day during fasting (**ST16, ST12**).

**Table 3.**
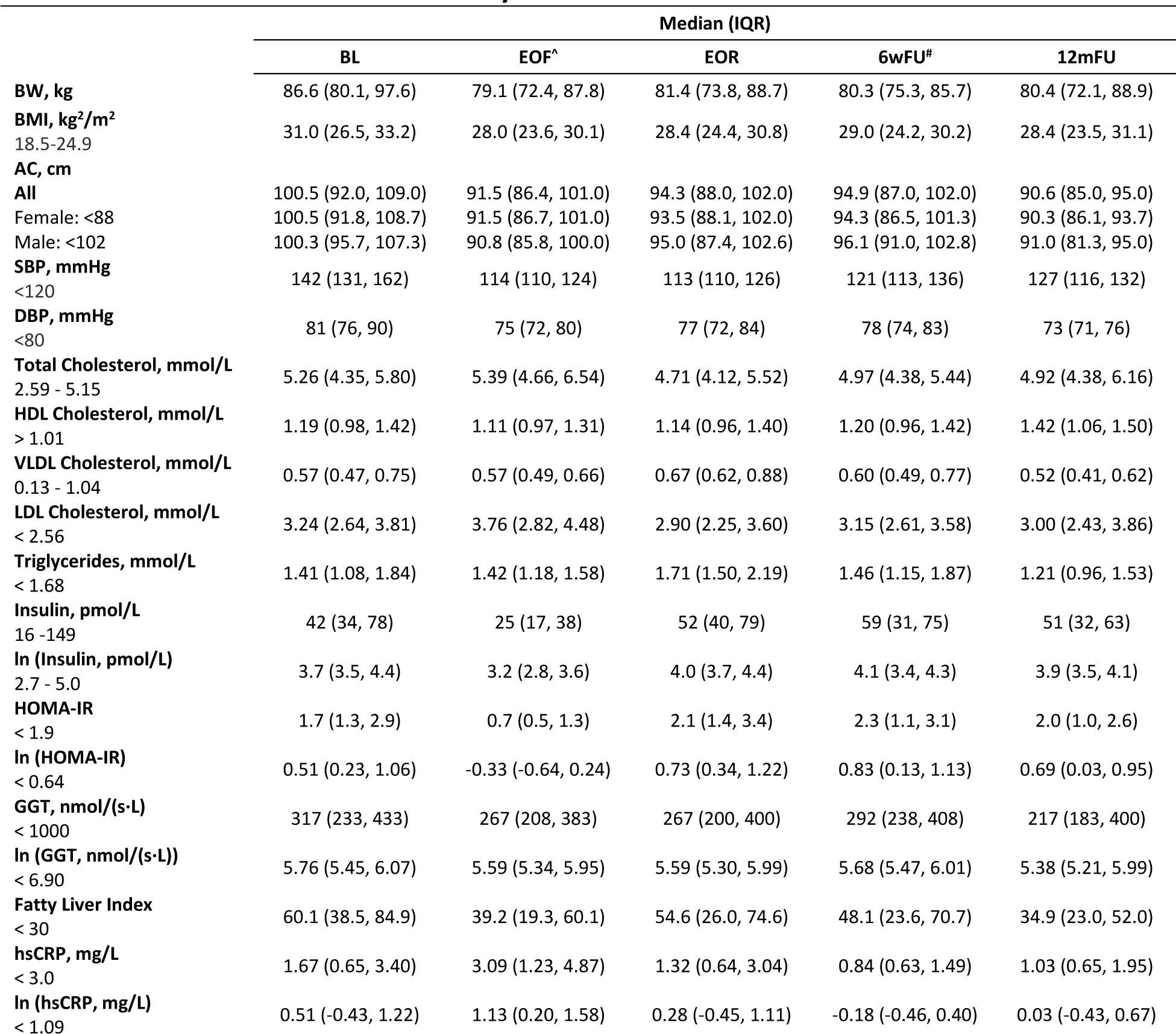
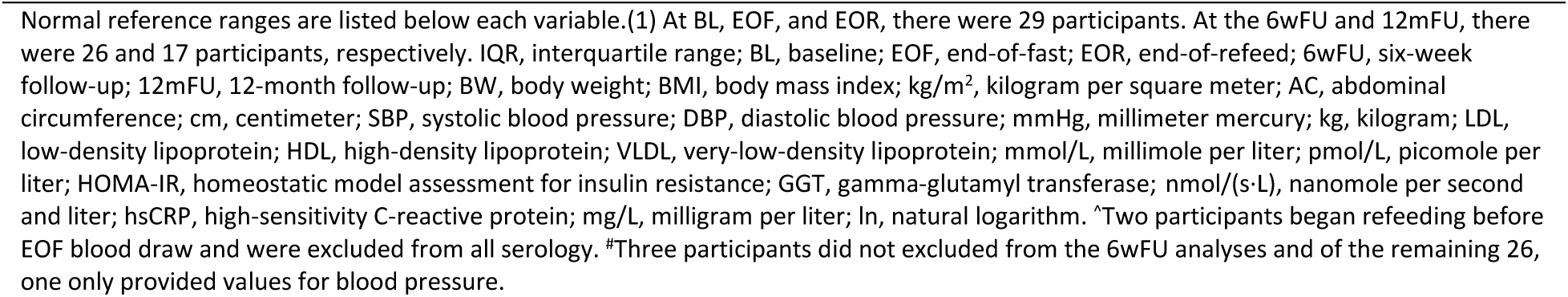
Cardiometabolic Biomarkers by Visit.

|  | Median (IQR) |  |  |  |  |
| --- | --- | --- | --- | --- | --- |
|  | BL | EOF<sup>a</sup> | EOR | 6wFU<sup>#</sup> | 12mFU |
| **BW, kg** | 86.6 (80.1, 97.6) | 79.1 (72.4, 87.8) | 81.4 (73.8, 88.7) | 80.3 (75.3, 85.7) | 80.4 (72.1, 88.9) |
| **BMI, kg<sup>2</sup>/m<sup>2</sup>**<br>18.5-24.9 | 31.0 (26.5, 33.2) | 28.0 (23.6, 30.1) | 28.4 (24.4, 30.8) | 29.0 (24.2, 30.2) | 28.4 (23.5, 31.1) |
| **AC, cm** |  |  |  |  |  |
| All | 100.5 (92.0, 109.0) | 91.5 (86.4, 101.0) | 94.3 (88.0, 102.0) | 94.9 (87.0, 102.0) | 90.6 (85.0, 95.0) |
| Female: <88 | 100.5 (91.8, 108.7) | 91.5 (86.7, 101.0) | 93.5 (88.1, 102.0) | 94.3 (86.5, 101.3) | 90.3 (86.1, 93.7) |
| Male: <102 | 100.3 (95.7, 107.3) | 90.8 (85.8, 100.0) | 95.0 (87.4, 102.6) | 96.1 (91.0, 102.8) | 91.0 (81.3, 95.0) |
| **SBP, mmHg**<br><120 | 142 (131, 162) | 114 (110, 124) | 113 (110, 126) | 121 (113, 136) | 127 (116, 132) |
| **DBP, mmHg**<br><80 | 81 (76, 90) | 75 (72, 80) | 77 (72, 84) | 78 (74, 83) | 73 (71, 76) |
| **Total Cholesterol, mmol/L**<br>2.59 - 5.15 | 5.26 (4.35, 5.80) | 5.39 (4.66, 6.54) | 4.71 (4.12, 5.52) | 4.97 (4.38, 5.44) | 4.92 (4.38, 6.16) |
| **HDL Cholesterol, mmol/L**<br>> 1.01 | 1.19 (0.98, 1.42) | 1.11 (0.97, 1.31) | 1.14 (0.96, 1.40) | 1.20 (0.96, 1.42) | 1.42 (1.06, 1.50) |
| **VLDL Cholesterol, mmol/L**<br>0.13 - 1.04 | 0.57 (0.47, 0.75) | 0.57 (0.49, 0.66) | 0.67 (0.62, 0.88) | 0.60 (0.49, 0.77) | 0.52 (0.41, 0.62) |
| **LDL Cholesterol, mmol/L**<br>< 2.56 | 3.24 (2.64, 3.81) | 3.76 (2.82, 4.48) | 2.90 (2.25, 3.60) | 3.15 (2.61, 3.58) | 3.00 (2.43, 3.86) |
| **Triglycerides, mmol/L**<br>< 1.68 | 1.41 (1.08, 1.84) | 1.42 (1.18, 1.58) | 1.71 (1.50, 2.19) | 1.46 (1.15, 1.87) | 1.21 (0.96, 1.53) |
| **Insulin, pmol/L**<br>16 -149 | 42 (34, 78) | 25 (17, 38) | 52 (40, 79) | 59 (31, 75) | 51 (32, 63) |
| **ln (Insulin, pmol/L)**<br>2.7 - 5.0 | 3.7 (3.5, 4.4) | 3.2 (2.8, 3.6) | 4.0 (3.7, 4.4) | 4.1 (3.4, 4.3) | 3.9 (3.5, 4.1) |
| **HOMA-IR**<br>< 1.9 | 1.7 (1.3, 2.9) | 0.7 (0.5, 1.3) | 2.1 (1.4, 3.4) | 2.3 (1.1, 3.1) | 2.0 (1.0, 2.6) |
| **ln (HOMA-IR)**<br>< 0.64 | 0.51 (0.23, 1.06) | -0.33 (-0.64, 0.24) | 0.73 (0.34, 1.22) | 0.83 (0.13, 1.13) | 0.69 (0.03, 0.95) |
| **GGT, nmol/(s·L)**<br>< 1000 | 317 (233, 433) | 267 (208, 383) | 267 (200, 400) | 292 (238, 408) | 217 (183, 400) |
| **ln (GGT, nmol/(s·L))**<br>< 6.90 | 5.76 (5.45, 6.07) | 5.59 (5.34, 5.95) | 5.59 (5.30, 5.99) | 5.68 (5.47, 6.01) | 5.38 (5.21, 5.99) |
| **Fatty Liver Index**<br>< 30 | 60.1 (38.5, 84.9) | 39.2 (19.3, 60.1) | 54.6 (26.0, 74.6) | 48.1 (23.6, 70.7) | 34.9 (23.0, 52.0) |
| **hsCRP, mg/L**<br>< 3.0 | 1.67 (0.65, 3.40) | 3.09 (1.23, 4.87) | 1.32 (0.64, 3.04) | 0.84 (0.63, 1.49) | 1.03 (0.65, 1.95) |
| **ln (hsCRP, mg/L)**<br>< 1.09 | 0.51 (-0.43, 1.22) | 1.13 (0.20, 1.58) | 0.28 (-0.45, 1.11) | -0.18 (-0.46, 0.40) | 0.03 (-0.43, 0.67) |

Normal reference ranges are listed below each variable.(1) At BL, EOF, and EOR, there were 29 participants. At the 6wFU and 12mFU, there were 26 and 17 participants, respectively. IQR, interquartile range; BL, baseline; EOF, end-of-fast; EOR, end-of-refeed; 6wFU, six-week follow-up; 12mFU, 12-month follow-up; BW, body weight; BMI, body mass index; kg/m<sup>2</sup>, kilogram per square meter; AC, abdominal circumference; cm, centimeter; SBP, systolic blood pressure; DBP, diastolic blood pressure; mmHg, millimeter mercury; kg, kilogram; LDL, low-density lipoprotein; HDL, high-density lipoprotein; VLDL, very-low-density lipoprotein; mmol/L, millimole per liter; pmol/L, picomole per liter; HOMA-IR, homeostatic model assessment for insulin resistance; GGT, gamma-glutamyl transferase; nmol/(s·L), nanomole per second and liter; hsCRP, high-sensitivity C-reactive protein; mg/L, milligram per liter; ln, natural logarithm. ^Two participants began refeeding before EOF blood draw and were excluded from all serology. \*Three participants did not excluded from the 6wFU analyses and of the remaining 26, one only provided values for blood pressure.

### Changes in SBP, DBP, and Anti-hypertensive Medication Use

During prefeeding and fasting, SBP reduced by an average of −9 mmHg/day and −1 mmHg/day, respectively (**ST12**, **SF3B**). DBP also significantly reduced during fasting by −0.3 mmHg per day (**ST12**, **SF3C**). Overall, baseline median (IQR) SBP/DBP reduced from 142/81 (131/76, 162/90) mmHg to 114/75 (110/72, 124/80) mmHg and 113/77 (110/72, 126/84) at EOF and EOR visits, respectively (**Table 3, ST13**). The significant reductions were sustained at the 6wFU and 12mFU visits. The estimated loss (95% CI) in SBP/DBP from the BL visit was −19.19/-5.13 (−26.32/-8.77, −12.03/-1.46) and −17.20/ −8.89 (−25.42/-13.09, −8.99/-4.67) mmHg at 6wFU and 12mFU visits, respectively (**ST13**).

At the BL visit, 15/29 (52%) participants were taking 19 anti-hypertensive medications; 11/15 were taking one and 4/15 were taking two medications (**ST17**). Only 5/15 (33%) had achieved a medication-controlled SBP/DBP of <130/80 mmHg (**Table 3**, **Figure 4**). During the prefeeding period, all participants were tapered completely off anti-hypertensive medications by a trained physician according to their individual medical needs and remained off medications while at the residential fasting center. Participants’ medication use after leaving the fasting center was determined independently by their primary care physician. Fourteen of the 15 participants who were taking anti-hypertensive medications at the BL visit returned for the 6wFU visit, of them only 1/14 was taking one anti-hypertensive medication at a reduced dose (**ST17**, **Figure 4**). Ten of the 15 participants who were taking anti-hypertensive medications at the BL visit returned for the 12mFU visit, of them 6/10 were taking anti-hypertensive medications (**ST17**, **Figure 4**). **Figure 4** shows the SBP/DBP distribution of medicated and unmedicated participants at each study visit.

### Changes in Select Cardiometabolic Markers

Total cholesterol, HDL, LDL, VLDL, triglycerides, hsCRP, insulin, GGT, as well as calculated HOMA-IR and FLI were also measured at each study visit (**Table 3**). At BL, only median total cholesterol, LDL, and FLI were outside of the normal limits. Median (IQR) total cholesterol normalized during refeeding to 4.71 (4.12, 5.52) mmol/L and was sustained at 6wFU and 12mFU visits. Median LDL remained slightly elevated throughout the study and median triglycerides increased slightly out of normal limits at the EOR visit but normalized again at the 6wFU visit. Median insulin remained within normal limits throughout the study but significantly increased at the EOR visit and normalized again by the 6wFU visit. Median (IQR) FLI, which was abnormally high at the BL visit, significantly decreased at 6wFU and 12mFU visits (**Table 3, ST13, Figure 4D**).

## 4 Discussion

These data contribute to a growing body of evidence suggesting that medically supervised prolonged water-only fasting followed by whole-plant-food refeeding may be a safe and feasible treatment option for various health conditions, particularly obesity and hypertension.[12; 13; 15; 36; 37] Importantly, the fasting and refeeding intervention in this study was conducted in a residential setting using a protocol in which participants had access to medical support 24 hours per day and are individually examined twice daily to identify potentially serious complications. Even though the prolonged intervention takes place in a residential setting, participants reported that they found it to be highly acceptable and preferable to conventional treatment with anti-hypertensive medications. Furthermore, the vast majority of participants were able to complete at least seven consecutive days of fasting, suggesting that the intervention is feasible.

There were no higher grade or serious adverse events identified in this study. Furthermore, the AE data reported in this hypertensive population are in agreement with previously reported retrospective data from a larger mixed normotensive and hypertensive population using the same fasting protocol.[14] Both studies identified that the most commonly reported AEs (occurring in >25% of participants) included transient mild or moderate fatigue, nausea, presyncope, dizziness, headache, and insomnia.[14] The present study also utilized hematology and serology to thoroughly screen for AEs and identified some commonly occurring AEs not previously reported. For example, 72% of participants experienced mild decreased blood bicarbonate while fasting potentially due to the overproduction of ketoacids.[38] Symptoms of decreased blood bicarbonate include fatigue and nausea, which could potentially explain, at least in part, the high rates of fatigue and nausea commonly reported while fasting.

HTN was also amongst the most commonly identified AEs in both studies. It is worth underscoring that fasting does not increase blood pressure in normotensive individuals.[39] Seven of the eight G3 events identified in this study were HTN and occurred during fasting, which is unsurprising given that this population had uncontrolled HTN and/or were weaned off anti-hypertensive medications before starting to fast. All of the G3 HTN events resolved while refeeding. The other G3 event was a low neutrophil count during refeeding that progressed from a G1 event during prefeeding. At the EOR visit, the participant’s WBC count had also decreased to a G1 event. Overall, we observed a trend of significantly decreased WBC count (**ST7**, **ST8**) at EOR, but the median value did not decrease below the lower normal level and abnormal values were not associated with any clinical outcomes. Previous research suggests that prolonged fasting induces stem cell regeneration and leads to a transient reduction in WBC count such that the body discards old WBCs before the immune system is replenished with new ones.[40] We do not know the relevance of this or other changes in CBC or CMP, but the results are in line with previous reports and warrant a more thorough analysis beyond the scope of this manuscript.[41]

Similar to previous reports, we observed clinically meaningful and significant reductions in median BP, BW, BMI, AC, and FLI that correlated with the intervention and were sustained at the 6wFU.[15] Results were also sustained at an unanticipated visit 12 months after the 6wFU visit. These preliminary data are confounded by low retention and that 5/17 participants underwent at least one other fasting and refeeding intervention during the study year. Nonetheless, the long-term BW loss data is especially encouraging given the long-held belief that weight lost through fasting is quickly regained.[42]

Although the study was not designed to test the effectiveness of fasting and refeeding in the treatment of hypertension, we observed that median unmedicated SBP reduced from Stage 2 HTN at the BL visit to normotensive by the EOR visit. At the 6wFU visit, only one participant had resumed anti-hypertensive medications and 38% of participants had sustained an SBP/DBP of <120/80 mmHg, with the largest reductions observed in participants who had stage 2 HTN at the BL visit. This large reduction was observed even though 5/17 participants with stage 2 HTN on enrollment continued to have stage 2 HTN six weeks after the intervention. The reductions in BP correlate with the timing of the intervention and are in agreement with other studies, [12; 13; 15; 36; 37] setting a precedent for future randomized controlled trials. Results from the 12mFU visit are inconclusive due to participant dropout, but the median SBP remained below 130 mmHg and of the 6/17 participants who resumed anti-hypertensive medications, only one had SBP below 120 mmHg.

These results are encouraging, but the study has several limitations that should be addressed in future research. A major limitation is the potential for selection bias since the small number of participants were recruited from existing patients at a single fasting center and the majority of enrolled participants were post-menopausal women. Therefore, the results from this “self-selected” population may not be representative of the general hypertensive population. The study also lacks a diet control group and/or usual care comparison group. Therefore, it is inconclusive as to what long-term results are attributable to diet alone and how the intervention compares to standard interventions (e.g., combined-therapy anti-hypertensive medications). Furthermore, the study was not powered to assess changes in cardiometabolic markers and only 59% of participants re-enrolled for the 12-month follow-up visit. Future studies should aim to enroll and randomize a sufficient number of participants from a more general hypertensive population and include appropriate control populations as well as hold more frequent, planned study visits. Another important methodological limitation is that BP data are based on single rather than 24-hour ambulatory BP measurements.[43] The effect of these methodological limitations on study outcomes is unknown, but these results are comparable to other reports in different populations.[12; 13; 36]

### Conclusion

Hypertension is the most common chronic disease worldwide, and when uncontrolled it is a risk factor for developing cardiovascular and other diseases. Standard medical care typically includes anti-hypertensive medications that only minimally lower overall health risk reductions, may be accompanied with serious side effects, have low adherence, and a high life-time cost.[4; 8; 44; 45] Our results suggest that prolonged water-only fasting followed by partial adherence to an SOS-free diet may be a safe and feasible alternative that results in sustained reductions in BMI and BP. These findings are in support of additional research to include a more diverse population, optimize treatment length and frequency, determine the effects of diet and other lifestyle modifications on sustained outcomes, and compare this intervention to standard treatment.

## Supplementary Materials

Supplemental Materials include **Supplemental Tables**: ST1. Enrollment and Visit Characteristics; ST2. Baseline ICD-10 Diagnosis & Code by Sex; ST3. Treatment Characteristics; ST4. SOS-Free Diet Screener Serving Quantity Reported by Category at BL, 6wFU, and 12mFU; ST5. Adverse Events Occurring in ≥ 10% of Participants by Treatment Stage and Grade; ST6. Total Adverse Events by Classification, Grade, and Outcome; ST7. CBC by Visit; ST8. Significance of Differences for CBC; ST9. CMP by Visit; ST10. Significance of Differences for CMP; ST11. 24-hour Dipstick Urinalysis by Visit; ST12. Daily Rate of Change for Body Weight, Blood Pressure, and Vital Signs; ST13. Significance of Difference for Cardiometabolic Biomarkers; ST14. Weight Class by Visit; ST15. Vital Signs by Visit; ST16. Significance of Differences for Vital Signs; ST17. Anti-Hypertensive Medication Use at BL, 6wFU, and 12mFU; **Supplemental Figures**: SF1: Percentage of participants responding to individual TAAS questions; SF2: Percentage of participants responding to individual FAQ questions; SF3: Daily measurements during prefeeding, fasting, and refeeding of BW (A), SBP (B), and DBP (C); SF4: BW (A), BMI (B), AC (C), and FLI (D) by visit.

## Non-standard Abbreviations and Acronyms

BL: Baseline
EOF: End of fast
EOR: End of refeed
FAQ: Food Acceptability Questionnaire
FU: Follow up
SOS: Salt, oil, and sugar
TAAS: Treatment Adherence/Acceptability Scale
VB: Vegetable Broth

## Acknowledgments

We would like to thank the dedicated staff at the TrueNorth Health Center, especially Justin Wise, Rie Schur, Ahmed Deen, Mara Gilbert, Faye Alexandrakis, and Haley Ramsey, for their contributions with participant monitoring, recording of adverse events, and REDCap instrument development. We also thank David Goldman for critically reviewing this manuscript.

## Author Contributions

Conceptualization, TRM, ES, and ACG; Methodology, TRM, ACG, MN; Formal Analysis, MN and EZ; Validation, MN; Investigation, SG, EZ, and NT; Resources, TRM, SG, and EZ; Data Curation, EZ, SG, and MN; Writing – Original Draft Preparation, EZ and TRM; Writing – Review & Editing, TRM, EZ, SG, MN, NT, ES, and A.C.G.; Visualization, EZ, MN, and TRM; Supervision, TRM; Project Administration, TRM, SG, and EZ; Funding Acquisition, ACG.

## Institutional Review Board Statement

The study was conducted according to the guidelines of the Declaration of Helsinki, and approved by the Institutional Review Board of the TrueNorth Health Foundation (TNHF2020-1HTN, August 16, 2020 and TNHF2021-1HTNFU, October 28, 2021).

## Informed Consent Statement

Informed consent was obtained from all subjects involved in the study.

## Data Availability Statement

Data is available upon request to the corresponding author.

## References

[1] Y. Ostchega, C.D. Fryar, T. Nwankwo, and D.T. Nguyen, Hypertension Prevalence Among Adults Aged 18 and Over: United States, 2017-2018. NCHS Data Brief (2020) 1-8.

[2] E. Oliveros, H. Patel, S. Kyung, S. Fugar, A. Goldberg, N. Madan, and K.A. Williams, Hypertension in older adults: Assessment, management, and challenges. Clin Cardiol 43 (2020) 99–107.

[3] S.S. Franklin, M.G. Larson, S.A. Khan, N.D. Wong, E.P. Leip, W.B. Kannel, and D. Levy, Does the relation of blood pressure to coronary heart disease risk change with aging? The Framingham Heart Study. Circulation 103 (2001) 1245–9.

[4] E.B. Kirkland, M. Heincelman, K.G. Bishu, S.O. Schumann, A. Schreiner, R.N. Axon, P.D. Mauldin, and W.P. Moran, Trends in Healthcare Expenditures Among US Adults With Hypertension: National Estimates, 2003-2014. J Am Heart Assoc 7 (2018).

[5] P.K. Whelton, R.M. Carey, W.S. Aronow, D.E. Casey, Jr., K.J. Collins, C. Dennison Himmelfarb, S.M. DePalma, S. Gidding, K.A. Jamerson, D.W. Jones, E.J. MacLaughlin, P. Muntner, B. Ovbiagele, S.C. Smith, Jr., C.C. Spencer, R.S. Stafford, S.J. Taler, R.J. Thomas, K.A. Williams, Sr., J.D. Williamson, and J.T. Wright, Jr., 2017 ACC/AHA/AAPA/ABC/ACPM/AGS/APhA/ASH/ASPC/NMA/PCNA Guideline for the Prevention, Detection, Evaluation, and Management of High Blood Pressure in Adults: A Report of the American College of Cardiology/American Heart Association Task Force on Clinical Practice Guidelines. J Am Coll Cardiol 71 (2018) e127-e248.

[6] M.D. Chobufo, V. Gayam, J. Soluny, E.U. Rahman, S. Enoru, J.B. Foryoung, V.N. Agbor, A. Dufresne, and T. Nfor, Prevalence and control rates of hypertension in the USA: 2017-2018. Int J Cardiol Hypertens 6 (2020) 100044.

[7] A. Albasri, M. Hattle, C. Koshiaris, A. Dunnigan, B. Paxton, S.E. Fox, M. Smith, L. Archer, B. Levis, R.A. Payne, R.D. Riley, N. Roberts, K.I.E. Snell, S. Lay-Flurrie, J. Usher-Smith, R. Stevens, F.D.R. Hobbs, R.J. McManus, J.P. Sheppard, and S. investigators, Association between antihypertensive treatment and adverse events: systematic review and meta-analysis. BMJ 372 (2021) n189.

[8] C.L.B. Ho, M. Breslin, J. Doust, C.M. Reid, and M.R. Nelson, Effectiveness of blood pressure-lowering drug treatment by levels of absolute risk: post hoc analysis of the Australian National Blood Pressure Study. BMJ Open 8 (2018) e017723.

[9] S.S. Virani, A. Alonso, H.J. Aparicio, E.J. Benjamin, M.S. Bittencourt, C.W. Callaway, A.P. Carson, A.M. Chamberlain, S. Cheng, F.N. Delling, M.S.V. Elkind, K.R. Evenson, J.F. Ferguson, D.K. Gupta, S.S. Khan, B.M. Kissela, K.L. Knutson, C.D. Lee, T.T. Lewis, J. Liu, M.S. Loop, P.L. Lutsey, J. Ma, J. Mackey, S.S. Martin, D.B. Matchar, M.E. Mussolino, S.D. Navaneethan, A.M. Perak, G.A. Roth, Z. Samad, G.M. Satou, E.B. Schroeder, S.H. Shah, C.M. Shay, A. Stokes, L.B. VanWagner, N.Y. Wang, C.W. Tsao, E. American Heart Association Council on, C. Prevention Statistics, and S. Stroke Statistics, Heart Disease and Stroke Statistics-2021 Update: A Report From the American Heart Association. Circulation 143 (2021) e254-e743.

[10] N.T. Artinian, G.F. Fletcher, D. Mozaffarian, P. Kris-Etherton, L. Van Horn, A.H. Lichtenstein, S. Kumanyika, W.E. Kraus, J.L. Fleg, N.S. Redeker, J.C. Meininger, J. Banks, E.M. Stuart-Shor, B.J. Fletcher, T.D. Miller, S. Hughes, L.T. Braun, L.A. Kopin, K. Berra, L.L. Hayman, L.J. Ewing, P.A. Ades, J.L. Durstine, N. Houston-Miller, L.E. Burke, and N. American Heart Association Prevention Committee of the Council on Cardiovascular, Interventions to promote physical activity and dietary lifestyle changes for cardiovascular risk factor reduction in adults: a scientific statement from the American Heart Association. Circulation 122 (2010) 406–41.

[11] A. Dhakal, C.T. K, and M. Neupane, Adherence to lifestyle modifications and its associated factors in hypertensive patients. J Clin Nurs 31 (2022) 2181–2188.

[12] A. Goldhamer, D. Lisle, B. Parpia, S.V. Anderson, and T.C. Campbell, Medically supervised water-only fasting in the treatment of hypertension. J Manipulative Physiol Ther 24 (2001) 335–9.

[13] A.C. Goldhamer, D.J. Lisle, P. Sultana, S.V. Anderson, B. Parpia, B. Hughes, and T.C. Campbell, Medically supervised water-only fasting in the treatment of borderline hypertension. J Altern Complement Med 8 (2002) 643–50.

[14] J.S. Finnell, B.C. Saul, A.C. Goldhamer, and T.R. Myers, Is fasting safe? A chart review of adverse events during medically supervised, water-only fasting. BMC Complement Altern Med 18 (2018) 67.

[15] S. Gabriel, M. Ncube, E. Zeiler, N. Thompson, M.C. Karlsen, D.M. Goldman, Z. Glavas, A. Beauchesne, E. Scharf, A.C. Goldhamer, and T.R. Myers, A Six-Week Follow-Up Study on the Sustained Effects of Prolonged Water-Only Fasting and Refeeding on Markers of Cardiometabolic Risk. Nutrients 14 (2022).

[16] T.R. Myers, B. Saul, M. Karlsen, A. Beauchesne, Z. Glavas, M. Ncube, R. Bradley, and A.C. Goldhamer, Potential Effects of Prolonged Water-Only Fasting Followed by a Whole-Plant-Food Diet on Salty and Sweet Taste Sensitivity and Perceived Intensity, Food Liking, and Dietary Intake. Cureus 14 (2022) e24689.

[17] I. Milosevic, H.C. Levy, G.M. Alcolado, and A.S. Radomsky, The Treatment Acceptability/Adherence Scale: Moving Beyond the Assessment of Treatment Effectiveness. Cogn Behav Ther 44 (2015) 456–69.

[18] A.D. Fruge, K.S. Smith, A.J. Riviere, W. Demark-Wahnefried, A.E. Arthur, W.M. Murrah, C.D. Morrow, R.D. Arnold, and K. Braxton-Lloyd, Primary Outcomes of a Randomized Controlled Crossover Trial to Explore the Effects of a High Chlorophyll Dietary Intervention to Reduce Colon Cancer Risk in Adults: The Meat and Three Greens (M3G) Feasibility Trial. Nutrients 11 (2019).

[19] P.A. Harris, R. Taylor, R. Thielke, J. Payne, N. Gonzalez, and J.G. Conde, Research electronic data capture (REDCap)--a metadata-driven methodology and workflow process for providing translational research informatics support. J Biomed Inform 42 (2009) 377–81.

[20] NIH, Common Terminology Criteria for Adverse Events (CTCAE), NIH.

[21] C.B. Weir, and A. Jan, BMI Classification Percentile And Cut Off Points, StatPearls, Treasure Island (FL), 2023.

[22] B. Hedblad, P. Nilsson, L. Janzon, and G. Berglund, Relation between insulin resistance and carotid intima-media thickness and stenosis in non-diabetic subjects. Results from a cross-sectional study in Malmo, Sweden. Diabet Med 17 (2000) 299–307.

[23] D.J. Cuthbertson, J. Koskinen, E. Brown, C.G. Magnussen, N. Hutri-Kahonen, M. Sabin, P. Tossavainen, E. Jokinen, T. Laitinen, J. Viikari, O.T. Raitakari, and M. Juonala, Fatty liver index predicts incident risk of prediabetes, type 2 diabetes and non-alcoholic fatty liver disease (NAFLD). Ann Med 53 (2021) 1256–1264.

[24] R.C. Team, R: A language and environment for statistical computing. R Foundation for Statistical Computing, Vienna, Austria, 2022.

[25] A. Kuznetsova, P.B. Brockhoff, and R.H.B. Christensen, lmerTest Package: Tests in Linear Mixed Effects Models. Journal of Statistical Software 82 (2017) 1–26.

[26] D. Bates, M. Mächler, B. Bolker, and S. Walker, Fitting Linear Mixed-Effects Models Using lme4. Journal of Statistical Software 67 (2015) 1–48.

[27] B. Bolker, D. Robinson, D. Menne, J. Gabry, P. Buerkner, C. Hua, W. Petry, J. Wiley, P. Kennedy, E. Szöcs, I. Patil, V. Arel-Bundock, B. Denney, and C. Brunson, broom.mixed: Tidying Methods for Mixed Models. (2022).

[28] J.F. Wiley, multilevelTools: Multilevel and Mixed Effects Model Diagnostics and Effect Sizes. (2020).

[29] M. Koller, robustlmm: An R Package for Robust Estimation of Linear Mixed-Effects Models. Journal of Statistical Software 75 (2016) 1–24.

[30] M. Koller, and W.A. Stahel, Robust Estimation of General Linear Mixed Effects Models. Robust and Multivariate Statistical Methods (2023).

[31] D. Lüdecke, A. Bartel, C. Schwemmer, C. Powell, A. Djalovski, and J. Titz, sjPlot: Data Visualization for Statistics in Social Science. R package version 2.8.15. (2023).

[32] S.T. Brookes, E. Whitely, M. Egger, G.D. Smith, P.A. Mulheran, and T.J. Peters, Subgroup analyses in randomized trials: risks of subgroup-specific analyses; power and sample size for the interaction test. J Clin Epidemiol 57 (2004) 229–36.

[33] D.J. O’Keefe, Brief Report: Post Hoc Power, Observed Power, A Priori Power, Retrospective Power, Prospective Power, Achieved Power: Sorting Out Appropriate Uses of Statistical Power Analyses. Communication Methods and Measures 1 (2010) 291–299.

[34] N.L. Bolger, and Jean-Philippe, Intensive longitudinal methods: An introduction to diary and experience sampling research. Guilford Press (2013).

[35] J. Pinheiro, D. Bates, S. DebRoy, D. Sarkar, S. Heisterkamp, B.V. Willigen, J. Ranke, and R.C. Team, nlme: Linear and Nonlinear Mixed Effects Models. (2023).

[36] A. Maifeld, H. Bartolomaeus, U. Lober, E.G. Avery, N. Steckhan, L. Marko, N. Wilck, I. Hamad, U. Susnjar, A. Mahler, C. Hohmann, C.Y. Chen, H. Cramer, G. Dobos, T.R. Lesker, T. Strowig, R. Dechend, D. Bzdok, M. Kleinewietfeld, A. Michalsen, D.N. Muller, and S.K. Forslund, Fasting alters the gut microbiome reducing blood pressure and body weight in metabolic syndrome patients. Nat Commun 12 (2021) 1970.

[37] C. Laurens, F. Grundler, A. Damiot, I. Chery, A.L. Le Maho, A. Zahariev, Y. Le Maho, A. Bergouignan, G. Gauquelin-Koch, C. Simon, S. Blanc, and F. Wilhelmi de Toledo, Is muscle and protein loss relevant in long-term fasting in healthy men? A prospective trial on physiological adaptations. J Cachexia Sarcopenia Muscle 12 (2021) 1690–1703.

[38] J.A. Kraut, and N.E. Madias, Metabolic acidosis: pathophysiology, diagnosis and management. Nat Rev Nephrol 6 (2010) 274–85.

[39] Z. Dai, H. Zhang, F. Wu, Y. Chen, C. Yang, H. Wang, X. Sui, Y. Guo, B. Xin, Z. Guo, J. Xiong, B. Wu, and Y. Li, Effects of 10-Day Complete Fasting on Physiological Homeostasis, Nutrition and Health Markers in Male Adults. Nutrients 14 (2022).

[40] C.W. Cheng, G.B. Adams, L. Perin, M. Wei, X. Zhou, B.S. Lam, S. Da Sacco, M. Mirisola, D.I. Quinn, T.B. Dorff, J.J. Kopchick, and V.D. Longo, Prolonged fasting reduces IGF-1/PKA to promote hematopoietic-stem-cell-based regeneration and reverse immunosuppression. Cell Stem Cell 14 (2014) 810–23.

[41] E. Ogłodek, and W. Pilis, Is Water-Only Fasting Safe? Global Advances in Integrative Medicine and Health (2021).

[42] M. Fanti, A. Mishra, V.D. Longo, and S. Brandhorst, Time-Restricted Eating, Intermittent Fasting, and Fasting-Mimicking Diets in Weight Loss. Curr Obes Rep 10 (2021) 70–80.

[43] C. Pena-Hernandez, K. Nugent, and M. Tuncel, Twenty-Four-Hour Ambulatory Blood Pressure Monitoring. J Prim Care Community Health 11 (2020) 2150132720940519.

[44] E.A. Kaiser, U. Lotze, and H.H. Schäfer, Increasing complexity: which drug class to choose for treatment of hypertension in the elderly? Clinical Interventions in Aging 2014 (2014) 459–475.

[45] E.A. Gebreyohannes, A.S. Bhagavathula, T.B. Abebe, Y.G. Tefera, and T.M. Abegaz, Adverse effects and non-adherence to antihypertensive medications in University of Gondar Comprehensive Specialized Hospital. Clin Hypertens 25 (2019) 1.

[46] LabCorp, Blood Specimens: Chemistry and Hematology.

