## Supplemental Data for "Prolonged Water-only Fasting is a Safe and Feasible Treatment Option for Managing Stage 1 and 2 Hypertension"

### Supplementary Materials

#### 1. Supplementary Tables

##### ST1. Enrollment and Visit Characteristics

| Participants | N |
|---|---|
| Enrolled | 30 |
| Ineligible | 1 |
| Completed BL visit | 29 |
| Completed EOF visit | 29* |
| Completed EOR visit | 29 |
| Completed 6wkFU visit | 27† |
| Completed 12mFU visit | 17 |

N, number of participants; BL, baseline; EOF, end-of-fast; EOR, end-of-refeed; 6wkFU, six-week follow-up; 12mFU, 12-month follow-up. *2/29 participants began refeeding before EOF blood draw and were excluded for serology and urinalysis. †2/27 provided incomplete data, of which 1/2 provided all data except for anthropometric measurements and 1/2 only provided blood pressure data.

##### ST2. Baseline ICD-10 Diagnosis & Code by Sex

| Diagnosis* | Code | Overall N = 29 | Female N = 19 | Male N = 10 |
|---|---|---|---|---|
| Mixed hyperlipidemia | **E78.2** | 10 (34%) | 7 (37%) | 3 (30%) |
| Hypothyroidism, unspecified | **E03.9** | 3 (10%) | 3 (16%) | 0 (0%) |
| Insomnia, unspecified | **G47.00** | 3 (10%) | 1 (5.3%) | 2 (20%) |
| Gastro-esophageal reflux disease without esophagitis | **K21.9** | 3 (10%) | 3 (16%) | 0 (0%) |
| Localized edema | **R60.0** | 3 (10%) | 2 (11%) | 1 (10%) |
| Prediabetes | **R73.03** | 3 (10%) | 3 (10%) | 0 (0%) |
| Leiomyoma of uterus, unspecified | **D25.9** | 2 (6.9%) | 2 (11%) | 0 (0%) |
| Hyperlipidemia, unspecified | **E78.5** | 2 (11%) | 2 (11%) | 0 (0%) |
| Major depressive disorder, recurrent, moderate | **F33.1** | 2 (6.9%) | 1 (5.3%) | 1 (10%) |
| Adjustment disorder with anxiety | **F43.22** | 2 (6.9%) | 2 (11%) | 0 (0%) |
| Obstructive sleep apnea | **G47.33** | 2 (6.9%) | 1 (5.3%) | 1 (10%) |
| Asymptomatic varicose veins of bilateral lower extremities | **I83.93** | 2 (6.9%) | 1 (5.3%) | 1 (10%) |
| Allergic rhinitis due to pollen | **J30.1** | 2 (6.9%) | 0 (0%) | 2 (20%) |
| Pain in left hip | **M25.552** | 2 (10%) | 2 (16%) | 0 (0%) |
| Other cervical disc degeneration, unspecified cervical region | **M50.30** | 2 (6.9%) | 1 (5.3%) | 1 (10%) |
| Other intervertebral disc degeneration, lumbar region | **M51.36** | 2 (6.9%) | 1 (5.3%) | 1 (10%) |
| Cervicalgia | **M54.2** | 2 (6.9%) | 2 (11%) | 0 (0%) |
| Low back pain | **M54.5** | 2 (6.9%) | 2 (11%) | 0 (0%) |
| Stress incontinence | **N39.3** | 2 (6.9%) | 2 (11%) | 0 (0%) |

| | | | | |
|---|---|---|---|---|
| Benign prostatic hyperplasia with lower urinary tract symptoms | **N40.1** | 2 (6.9%) | 0 (0%) | 2 (20%) |
| Menopausal and female climacteric states | **N95.1** | 2 (6.9%) | 2 (11%) | 0 (0%) |
| Rash and other nonspecific skin eruption | **R21** | 2 (6.9%) | 2 (11%) | 0 (0%) |
| Nocturia | **R35.1** | 2 (6.9%) | 0 (0%) | 2 (20%) |
| Basal cell carcinoma of skin, unspecified | **C44.91** | 1 (3.4%) | 0 (0%) | 1 (10%) |
| Anemia, unspecified | **D64.9** | 1 (3.4%) | 1 (5.3%) | 0 (0%) |
| Nontoxic multinodular goiter | **E04.2** | 1 (3.4%) | 1 (5.3%) | 0 (0%) |
| Thyroiditis | **E06** | 1 (3.4%) | 1 (5.3%) | 0 (0%) |
| Type 2 diabetes mellitus with hyperglycemia | **E11.65** | 1 (3.4%) | 1 (5.3%) | 0 (0%) |
| Testicular hypofunction | **E29.1** | 1 (3.4%) | 0 (0%) | 1 (10%) |
| Alcohol dependence with alcohol-induced sleep disorder | **F10.282** | 1 (3.4%) | 1 (5.3%) | 0 (0%) |
| Post-traumatic stress disorder, chronic | **F43.12** | 1 (3.4%) | 1 (5.3%) | 0 (0%) |
| Migraine, unspecified, not intractable, without status migrainosus | **G43.909** | 1 (3.4%) | 1 (5.3%) | 0 (0%) |
| Episodic cluster headache, not intractable | **G44.019** | 1 (3.4%) | 1 (5.3%) | 0 (0%) |
| Unspecified glaucoma | **H40.9** | 1 (3.4%) | 1 (5.3%) | 0 (0%) |
| Left bundle-branch block, unspecified | **I44.7** | 1 (3.4%) | 1 (5.3%) | 0 (0%) |
| Cardiac arrhythmia, unspecified | **I49.9** | 1 (3.4%) | 1 (5.3%) | 0 (0%) |
| Varicose veins of unspecified lower extremity with inflammation | **I83.10** | 1 (3.4%) | 1 (5.3%) | 0 (0%) |
| Allergic rhinitis, unspecified | **J30.9** | 1 (3.4%) | 1 (5.3%) | 0 (0%) |
| Unspecified asthma, uncomplicated | **J45.909** | 1 (3.4%) | 1 (5.3%) | 0 (0%) |
| Diaphragmatic hernia without obstruction or gangrene | **K44.9** | 1 (3.4%) | 1 (5.3%) | 0 (0%) |
| Rectal prolapse | **K62.3** | 1 (3.4%) | 1 (5.3%) | 0 (0%) |
| Polyp of colon | **K63.5** | 1 (3.4%) | 1 (5.3%) | 0 (0%) |
| Calculus of gallbladder without cholecystitis without obstruction | **K80.20** | 1 (3.4%) | 0 (0%) | 1 (10%) |
| Local infection of the skin and subcutaneous tissue, unspecified | **L08.9** | 1 (3.4%) | 1 (5.3%) | 0 (0%) |
| Dermatitis, unspecified | **L30.9** | 1 (3.4%) | 1 (5.3%) | 0 (0%) |
| Lichen sclerosus et atrophicus | **L90.0** | 1 (3.4%) | 1 (5.3%) | 0 (0%) |
| Bilateral primary osteoarthritis of knee | **M17.0** | 1 (3.4%) | 1 (5.3%) | 0 (0%) |
| Unspecified osteoarthritis, unspecified site | **M19.90** | 1 (3.4%) | 1 (5.3%) | 0 (0%) |
| Pain in unspecified joint | **M25.50** | 1 (3.4%) | 1 (5.3%) | 0 (0%) |

| | | | | |
|---|---|---|---|---|
| Pain in right shoulder | **M25.511** | 1 (3.4%) | 0 (0%) | 1 (10%) |
| Pain in right hip | **M25.551** | 1 (6.9%) | 1 (11%) | 0 (0%) |
| Pain in unspecified hip | **M25.559** | 1 (3.4%) | 0 (0%) | 1 (10%) |
| Pain in right knee | **M25.561** | 1 (6.9%) | 1 (11%) | 0 (0%) |
| Pain in right hand | **M79.641** | 1 (3.4%) | 0 (0%) | 1 (10%) |
| Pain in left hand | **M79.642** | 1 (3.4%) | 0 (0%) | 1 (10%) |
| Age-related osteoporosis without current pathological fracture | **M81.0** | 1 (3.4%) | 1 (5.3%) | 0 (0%) |
| Segmental and somatic dysfunction of cervical region | **M99.01** | 1 (6.9%) | 0 (0%) | 1 (10%) |
| Segmental and somatic dysfunction of thoracic region | **M99.02** | 1 (11%) | 1 (11%) | 0 (0%) |
| Segmental and somatic dysfunction of sacral region | **M99.04** | 1 (11%) | 1 (11%) | 0 (0%) |
| Uterovaginal prolapse, unspecified | **N81.4** | 1 (3.4%) | 1 (5.3%) | 0 (0%) |
| Palpitations | **R00.2** | 1 (3.4%) | 1 (5.3%) | 0 (0%) |
| Cardiac murmur, unspecified | **R01.1** | 1 (3.4%) | 1 (5.3%) | 0 (0%) |
| Abdominal distension (gaseous) | **R14.0** | 1 (3.4%) | 1 (5.3%) | 0 (0%) |
| Inappropriate diet and eating habits | **Z72.4** | 1 (3.4%) | 1 (5.3%) | 0 (0%) |
| Hormone replacement therapy | **Z79.890** | 1 (3.4%) | 1 (5.3%) | 0 (0%) |
| Personal history of malignant neoplasm of brain | **Z85.841** | 1 (3.4%) | 1 (5.3%) | 0 (0%) |
| Personal history of other diseases of the musculoskeletal system and connective tissue | **Z87.39** | 1 (3.4%) | 0 (0%) | 1 (10%) |
| Personal history of urinary calculi | **Z87.442** | 1 (3.4%) | 0 (0%) | 1 (10%) |
| Presence of aortocoronary bypass graft | **Z95.1** | 1 (3.4%) | 1 (5.3%) | 0 (0%) |
| Presence of coronary angioplasty implant and graft | **Z95.5** | 1 (3.4%) | 1 (5.3%) | 0 (0%) |
| Presence of right artificial knee joint | **Z96.651** | 1 (3.4%) | 0 (0%) | 1 (10%) |

Participants may have had more than one diagnoses. ICD, international classification of disease;[1] N, total number; %, percent. *Preexisting conditions, not including hypertension.

**ST3. Treatment Characteristics**

| **Duration, d** | **Median (Range)** |
|---|---|
| Prefeeding Period | 2 (1, 4) |
| Fasting Period | 11 (7, 40) |
| Refeeding Period | 5 (3, 17) |
| 6wkFU Period | 45 (40, 58) |
| 12mFU Period | 364 (358, 383) |
| **Type** | **N (%)** |
| Water Only | 17 (58.6%) |
| Water & VB | 9 (31.0%) |

| | |
|---|---|
| VB Only | 1 (3.4%) |
| Water & Juice | 1 (3.4%) |
| Water & P/Z blend | 1 (3.4%) |
| **Additional Treatments* between 6wkFU and 12mFU** | **N (%)** |
| No Treatment | 12 (71%) |
| 1 Treatment | 2 (12%) |
| 2 Treatments | 3 (18%) |

d, day; 6wkFU, six-week follow-up; 12mFU, 12-month follow-up; VB, vegetable broth; P/Z, potato/zucchini. Due to insignificant amounts of calories in VB it is assumed equivalent to water-only fasting. *Of the eight additional treatments, seven were water-only and one was a combination of water-only and juice with a median (range) fasting and refeeding length of 10 (8, 13) and 6 (4, 9) days, respectively.

#### ST4. SOS-Free Diet Screener Serving Quantity Reported by Category at BL, 6wkFU, and 12mFU

| | N (%) | | | | | | | | | | | |
|---|---|---|---|---|---|---|---|---|---|---|---|---|
| **Number of Servings** | **0 per month** | **1 per month** | **2-3 per month** | **1 per week** | **2 per week** | **3-4 per week** | **5-6 per week** | **1 per day** | **2 per day** | **3 per day** | **4 per day** | **≥ 5 per day** |
| **Vegetables** | | | | | | | | | | | | |
| **BL** | 0 (0%) | 0 (0%) | 0 (0%) | 0 (0%) | 0 (0%) | 4 (14%) | 0 (0%) | 5 (17%) | 4 (14%) | 6 (21%) | 4 (14%) | 6 (21%) |
| **6wkFU** | 0 (0%) | 0 (0%) | 0 (0%) | 0 (0%) | 0 (0%) | 1 (4%) | 0 (0%) | 2 (8%) | 4 (15%) | 5 (19%) | 6 (23%) | 8 (31%) |
| **12mFU** | 0 (0%) | 0 (0%) | 0 (0%) | 0 (0%) | 0 (0%) | 1 (5.9%) | 1 (5.9%) | 0 (0%) | 4 (24%) | 2 (12%) | 4 (24%) | 5 (29%) |
| **Fruits** | | | | | | | | | | | | |
| **BL** | 0 (0%) | 1 (3%) | 2 (7%) | 2 (7%) | 0 (0%) | 2 (7%) | 0 (0%) | 2 (7%) | 9 (31%) | 5 (17%) | 4 (14%) | 2 (7%) |
| **6wkFU** | 0 (0%) | 0 (0%) | 0 (0%) | 0 (0%) | 2 (8%) | 0 (0%) | 0 (0%) | 4 (15%) | 4 (15%) | 9 (35%) | 5 (19%) | 2 (8%) |
| **12mFU** | 0 (0%) | 0 (0%) | 0 (0%) | 0 (0%) | 0 (0%) | 1 (5.9%) | 0 (0%) | 2 (12%) | 5 (29%) | 4 (24%) | 1 (5.9%) | 4 (24%) |
| **Avocado** | | | | | | | | | | | | |
| **BL** | 2 (7%) | 3 (10%) | 5 (17%) | 3 (10%) | 2 (7%) | 8 (28%) | 2 (7%) | 4 (14%) | 0 (0%) | 0 (0%) | 0 (0%) | 0 (0%) |
| **6wkFU** | 4 (15%) | 2 (8%) | 2 (8%) | 2 (8%) | 4 (15%) | 4 (15%) | 2 (8%) | 6 (23%) | 0 (0%) | 0 (0%) | 0 (0%) | 0 (0%) |
| **12mFU** | 2 (12%) | 4 (24%) | 3 (18%) | 0 (0%) | 2 (12%) | 4 (24%) | 1 (5.9%) | 1 (5.9%) | 0 (0%) | 0 (0%) | 0 (0%) | 0 (0%) |
| **Whole Grain** | | | | | | | | | | | | |
| **BL** | 1 (3%) | 1 (3%) | 3 (10%) | 3 (10%) | 1 (3%) | 6 (21%) | 3 (10%) | 4 (14%) | 5 (17%) | 1 (3%) | 0 (0%) | 0 (0%) |
| **6wkFU** | 0 (0%) | 0 (0%) | 0 (0%) | 0 (0%) | 2 (8%) | 5 (19%) | 3 (12%) | 6 (23%) | 6 (23%) | 2 (8%) | 2 (8%) | 0 (0%) |
| **12mFU** | 0 (0%) | 0 (0%) | 2 (12%) | 0 (0%) | 1 (5.9%) | 3 (18%) | 1 (5.9%) | 3 (18%) | 4 (24%) | 0 (0%) | 3 (18%) | 0 (0%) |
| **Whole Grain Flour** | | | | | | | | | | | | |
| **BL** | 6 (21%) | 1 (3%) | 1 (3%) | 3 (10%) | 7 (24%) | 7 (24%) | 1 (3%) | 2 (7%) | 1 (3%) | 0 (0%) | 0 (0%) | 0 (0%) |
| **6wkFU** | 4 (15%) | 2 (8%) | 3 (12%) | 2 (8%) | 7 (27%) | 4 (15%) | 1 (4%) | 3 (12%) | 0 (0%) | 0 (0%) | 0 (0%) | 0 (0%) |
| **12mFU** | 1 (5.9%) | 2 (12%) | 3 (18%) | 1 (5.9%) | 3 (18%) | 4 (24%) | 1 (5.9%) | 1 (5.9%) | 1 (5.9%) | 0 (0%) | 0 (0%) | 0 (0%) |
| **Refined Grain** | | | | | | | | | | | | |
| **BL** | 7 (24%) | 0 (0%) | 5 (17%) | 0 (0%) | 3 (10%) | 6 (21%) | 3 (10%) | 0 (0%) | 0 (0%) | 5 (17%) | 0 (0%) | 0 (0%) |
| **6wkFU** | 9 (35%) | 4 (15%) | 5 (19%) | 2 (8%) | 5 (19%) | 0 (0%) | 0 (0%) | 0 (0%) | 0 (0%) | 1 (4%) | 0 (0%) | 0 (0%) |
| **12mFU** | 3 (18%) | 2 (12%) | 1 (5.9%) | 3 (18%) | 3 (18%) | 3 (18%) | 0 (0%) | 0 (0%) | 2 (12%) | 0 (0%) | 0 (0%) | 0 (0%) |
| **Legumes** | | | | | | | | | | | | |
| **BL** | 1 (3%) | 0 (0%) | 2 (7%) | 1 (3%) | 2 (7%) | 8 (28%) | 4 (14%) | 2 (7%) | 5 (17%) | 2 (7%) | 2 (7%) | 0 (0%) |

| | | | | | | | | | | | | |
|---|---|---|---|---|---|---|---|---|---|---|---|---|
| **6wkFU** | 0 (0%) | 0 (0%) | 0 (0%) | 0 (0%) | 2 (8%) | 6 (23%) | 6 (23%) | 6 (23%) | 2 (8%) | 3 (12%) | 1 (4%) | 0 (0%) |
| **12mFU** | 0 (0%) | 0 (0%) | 0 (0%) | 1 (5.9%) | 2 (12%) | 3 (18%) | 2 (12%) | 4 (24%) | 3 (18%) | 1 (5.9%) | 0 (0%) | 1 (5.9%) |
| **Nuts and Seeds** | | | | | | | | | | | | |
| **BL** | 1 (3%) | 2 (7%) | 3 (10%) | 2 (7%) | 0 (0%) | 2 (7%) | 3 (10%) | 7 (24%) | 6 (21%) | 3 (10%) | 0 (0%) | 0 (0%) |
| **6wkFU** | 2 (8%) | 0 (0%) | 0 (0%) | 1 (4%) | 1 (4%) | 7 (27%) | 2 (8%) | 5 (19%) | 8 (31%) | 0 (0%) | 0 (0%) | 0 (0%) |
| **12mFU** | 1 (5.9%) | 1 (5.9%) | 1 (5.9%) | 0 (0%) | 1 (5.9%) | 2 (12%) | 0 (0%) | 2 (12%) | 7 (41%) | 2 (12%) | | |
| **Red Meat** | | | | | | | | | | | | |
| **BL** | 15 (52%) | 3 (10%) | 3 (10%) | 2 (7%) | 3 (10%) | 2 (7%) | 1 (3%) | 0 (0%) | 0 (0%) | 0 (0%) | 0 (0%) | 0 (0%) |
| **6wkFU** | 21 (81%) | 1 (4%) | 0 (0%) | 2 (8%) | 1 (4%) | 0 (0%) | 0 (0%) | 1 (4%) | 0 (0%) | 0 (0%) | 0 (0%) | 0 (0%) |
| **12mFU** | 13 (76%) | 0 (0%) | 1 (5.9%) | 2 (12%) | 0 (0%) | 1 (5.9%) | 0 (0%) | 0 (0%) | 0 (0%) | 0 (0%) | 0 (0%) | 0 (0%) |
| **Poultry** | | | | | | | | | | | | |
| **BL** | 17 (59%) | 0 (0%) | 1 (3%) | 4 (14%) | 5 (17%) | 1 (3%) | 1 (3%) | 0 (0%) | 0 (0%) | 0 (0%) | 0 (0%) | 0 (0%) |
| **6wkFU** | 22 (85%) | 1 (4%) | 1 (4%) | 2 (8%) | 0 (0%) | 0 (0%) | 0 (0%) | 0 (0%) | 0 (0%) | 0 (0%) | 0 (0%) | 0 (0%) |
| **12mFU** | 12 (71%) | 1 (5.9%) | 1 (5.9%) | 0 (0%) | 2 (12%) | 1 (5.9%) | 0 (0%) | 0 (0%) | 0 (0%) | 0 (0%) | 0 (0%) | 0 (0%) |
| **Fish** | | | | | | | | | | | | |
| **BL** | 10 (34%) | 7 (24%) | 5 (17%) | 2 (7%) | 2 (7%) | 3 (10%) | 0 (0%) | 0 (0%) | 0 (0%) | 0 (0%) | 0 (0%) | 0 (0%) |
| **6wkFU** | 19 (73%) | 3 (12%) | 2 (8%) | 1 (4%) | 1 (4%) | 0 (0%) | 0 (0%) | 0 (0%) | 0 (0%) | 0 (0%) | 0 (0%) | 0 (0%) |
| **12mFU** | 12 (71%) | 2 (12%) | 2 (12%) | 0 (0%) | 1 (5.9%) | 0 (0%) | 0 (0%) | 0 (0%) | 0 (0%) | 0 (0%) | 0 (0%) | 0 (0%) |
| **Dairy** | | | | | | | | | | | | |
| **BL** | 14 (48%) | 0 (0%) | 1 (3%) | 1 (3%) | 2 (7%) | 2 (7%) | 1 (3%) | 3 (10%) | 2 (7%) | 3 (10%) | 0 (0%) | 0 (0%) |
| **6wkFU** | 16 (62%) | 4 (15%) | 2 (8%) | 1 (4%) | 0 (0%) | 2 (8%) | 0 (0%) | 1 (4%) | 0 (0%) | 0 (0%) | 0 (0%) | 0 (0%) |
| **12mFU** | 12 (71%) | 2 (12%) | 1 (5.9%) | 0 (0%) | 0 (0%) | 1 (5.9%) | 1 (5.9%) | 0 (0%) | 0 (0%) | 0 (0%) | 0 (0%) | 0 (0%) |
| **Eggs** | | | | | | | | | | | | |
| **BL** | 13 (45%) | 2 (7%) | 2 (7%) | 2 (7%) | 4 (14%) | 3 (10%) | 2 (7%) | 1 (3%) | 0 (0%) | 0 (0%) | 0 (0%) | 0 (0%) |
| **6wkFU** | 22 (85%) | 0 (0%) | 2 (8%) | 1 (4%) | 1 (4%) | 0 (0%) | 0 (0%) | 0 (0%) | 0 (0%) | 0 (0%) | 0 (0%) | 0 (0%) |
| **12mFU** | 12 (71%) | 3 (18%) | 0 (0%) | 1 (5.9%) | 0 (0%) | 1 (5.9%) | 0 (0%) | 0 (0%) | 0 (0%) | 0 (0%) | 0 (0%) | 0 (0%) |
| **Prepared Food with Salt** | | | | | | | | | | | | |
| **BL** | 3 (10%) | 1 (3%) | 0 (0%) | 5 (17%) | 3 (10%) | 6 (21%) | 4 (14%) | 1 (3%) | 2 (7%) | 1 (3%) | 1 (3%) | 2 (7%) |
| **6wkFU** | 6 (23%) | 3 (12%) | 7 (27%) | 2 (8%) | 3 (12%) | 3 (12%) | 1 (4%) | 0 (0%) | 0 (0%) | 1 (4%) | 0 (0%) | 0 (0%) |
| **12mFU** | 2 (12%) | 2 (12%) | 2 (12%) | 3 (18%) | 4 (24%) | 2 (12%) | 0 (0%) | 1 (5.9%) | 1 (5.9%) | 0 (0%) | 0 (0%) | 0 (0%) |
| **Added Salt** | | | | | | | | | | | | |
| **BL** | 8 (28%) | 2 (7%) | 2 (7%) | 1 (3%) | 3 (10%) | 3 (10%) | 2 (7%) | 2 (7%) | 0 (0%) | 1 (3%) | 1 (3%) | 4 (14%) |
| **6wkFU** | 16 (62%) | 1 (4%) | 3 (12%) | 1 (4%) | 1 (4%) | 2 (8%) | 1 (4%) | 1 (4%) | 0 (0%) | 0 (0%) | 0 (0%) | 0 (0%) |
| **12mFU** | 7 (41%) | 1 (5.9%) | 1 (5.9%) | 3 (18%) | 2 (12%) | 0 (0%) | 1 (5.9%) | 2 (12%) | 0 (0%) | 0 (0%) | 0 (0%) | 0 (0%) |
| **Cooked with Salt** | | | | | | | | | | | | |
| **BL** | 7 (24%) | 2 (7%) | 1 (3%) | 0 (0%) | 4 (14%) | 5 (17%) | 2 (7%) | 1 (3%) | 3 (10%) | 1 (3%) | 0 (0%) | 3 (10%) |
| **6wkFU** | 12 (46%) | 5 (19%) | 2 (8%) | 2 (8%) | 0 (0%) | 1 (4%) | 1 (4%) | 2 (8%) | 0 (0%) | 1 (4%) | 0 (0%) | 0 (0%) |
| **12mFU** | 8 (47%) | 1 (5.9%) | 3 (18%) | 0 (0%) | 0 (0%) | 2 (12%) | 2 (12%) | 1 (5.9%) | 0 (0%) | 0 (0%) | 0 (0%) | 0 (0%) |
| **Prepared Food with Oil** | | | | | | | | | | | | |
| **BL** | 3 (10%) | 1 (3%) | 5 (17%) | 4 (14%) | 0 (0%) | 0 (0%) | 10 (34%) | 1 (3%) | 4 (14%) | 1 (3%) | 0 (0%) | 0 (0%) |

| | | | | | | | | | | | | |
|---|---|---|---|---|---|---|---|---|---|---|---|---|
| **6wkFU** | 9 (35%) | 6 (23%) | 6 (23%) | 2 (8%) | 0 (0%) | 0 (0%) | 1 (4%) | 1 (4%) | 1 (4%) | 0 (0%) | 0 (0%) | 0 (0%) |
| **12mFU** | 3 (18%) | 4 (24%) | 2 (12%) | 6 (35%) | 0 (0%) | 0 (0%) | 1 (5.9%) | 1 (5.9%) | 0 (0%) | 0 (0%) | 0 (0%) | 0 (0%) |
| **Added Oil** | | | | | | | | | | | | |
| **BL** | 16 (55%) | 1 (3%) | 4 (14%) | 0 (0%) | 1 (3%) | 1 (3%) | 1 (3%) | 3 (10%) | 0 (0%) | 1 (3%) | 1 (3%) | 0 (0%) |
| **6wkFU** | 19 (73%) | 3 (12%) | 1 (4%) | 2 (8%) | 1 (4%) | 0 (0%) | 0 (0%) | 0 (0%) | 0 (0%) | 0 (0%) | 0 (0%) | 0 (0%) |
| **12mFU** | 14 (82%) | 0 (0%) | 0 (0%) | 0 (0%) | 1 (5.9%) | 2 (12%) | 0 (0%) | 0 (0%) | 0 (0%) | 0 (0%) | 0 (0%) | 0 (0%) |
| **Cooked with Oil** | | | | | | | | | | | | |
| **BL** | 9 (31%) | 2 (7%) | 4 (14%) | 1 (3%) | 1 (3%) | 2 (7%) | 3 (10%) | 3 (10%) | 0 (0%) | 4 (14%) | 0 (0%) | 0 (0%) |
| **6wkFU** | 17 (65%) | 1 (4%) | 5 (19%) | 0 (0%) | 1 (4%) | 1 (4%) | 0 (0%) | 0 (0%) | 1 (4%) | 0 (0%) | 0 (0%) | 0 (0%) |
| **12mFU** | 9 (53%) | 1 (5.9%) | 0 (0%) | 3 (18%) | 1 (5.9%) | 3 (18%) | 0 (0%) | 0 (0%) | 0 (0%) | 0 (0%) | 0 (0%) | 0 (0%) |
| **Prepared Food with Added Sugar** | | | | | | | | | | | | |
| **BL** | 6 (21%) | 1 (3%) | 3 (10%) | 1 (3%) | 2 (7%) | 6 (21%) | 2 (7%) | 3 (10%) | 3 (10%) | 1 (3%) | 0 (0%) | 1 (3%) |
| **6wkFU** | 12 (50%) | 3 (12%) | 4 (17%) | 1 (4.2%) | 0 (0%) | 2 (8%) | 0 (0%) | 2 (8%) | 0 (0%) | 0 (0%) | 0 (0%) | 0 (0%) |
| **12mFU** | 4 (25%) | 2 (12%) | 3 (19%) | 2 (12%) | 1 (6.2%) | 1 (6.2%) | 0 (0%) | 0 (0%) | 2 (12%) | 1 (6.2%) | 0 (0%) | 0 (0%) |
| **Added Sugar** | | | | | | | | | | | | |
| **BL** | 18 (62%) | 1 (3%) | 2 (7%) | 1 (3%) | 1 (3%) | 0 (0%) | 2 (7%) | 3 (10%) | 0 (0%) | 0 (0%) | 0 (0%) | 1 (3%) |
| **6wkFU** | 24 (92%) | 0 (0%) | 2 (8%) | 0 (0%) | 0 (0%) | 0 (0%) | 0 (0%) | 0 (0%) | 0 (0%) | 0 (0%) | 0 (0%) | 0 (0%) |
| **12mFU** | 13 (76%) | 1 (5.9%) | 1 (5.9%) | 0 (0%) | 0 (0%) | 0 (0%) | 1 (5.9%) | 1 (5.9%) | 0 (0%) | 0 (0%) | 0 (0%) | 0 (0%) |
| **Cooked with Added Sugar** | | | | | | | | | | | | |
| **BL** | 11 (38%) | 1 (3%) | 6 (21%) | 3 (10%) | 2 (7%) | 1 (3%) | 2 (7%) | 2 (7%) | 0 (0%) | 1 (3%) | 0 (0%) | 0 (0%) |
| **6wkFU** | 21 (81%) | 2 (8%) | 2 (8%) | 0 (0%) | 0 (0%) | 1 (4%) | 0 (0%) | 0 (0%) | 0 (0%) | 0 (0%) | 0 (0%) | 0 (0%) |
| **12mFU** | 9 (53%) | 1 (5.9%) | 1 (5.9%) | 0 (0%) | 0 (0%) | 5 (29%) | 0 (0%) | 1 (5.9%) | 0 (0%) | 0 (0%) | 0 (0%) | 0 (0%) |
| **B12** | | | | | | | | | | | | |
| **BL** | 6 (21%) | 1 (3%) | 0 (0%) | 2 (7%) | 1 (3%) | 7 (24%) | 0 (0%) | 12 (41%) | 0 (0%) | 0 (0%) | 0 (0%) | 0 (0%) |
| **6wkFU** | 3 (12%) | 0 (0%) | 3 (12%) | 0 (0%) | 2 (8%) | 5 (19%) | 2 (8%) | 11 (42%) | 0 (0%) | 0 (0%) | 0 (0%) | 0 (0%) |
| **12mFU** | 3 (18%) | 0 (0%) | 0 (0%) | 3 (18%) | 0 (0%) | 4 (24%) | 0 (0%) | 7 (41%) | 0 (0%) | 0 (0%) | 0 (0%) | 0 (0%) |

| **Additional Supplements** | | | | | | |
|---|---|---|---|---|---|---|
| | **None** | **Omega-3** | **Vitamin D** | **Iodine** | **Probiotics** | **Other** |
| **BL** | 5 (17%) | 6 (21%) | 23 (79%) | 0 (0%) | 4 (14%) | 9 (31%) |
| **6wkFU** | 7 (27%) | 4 (15%) | 18 (69%) | 2 (7.4%) | 3 (12%) | 6 (23%) |
| **12mFU** | 4 (24%) | 7 (41%) | 12 (71%) | 2 (12%) | 5 (29%) | 5 (29%) |

| | | | | | | | | | | | | |
|---|---|---|---|---|---|---|---|---|---|---|---|---|
| **Alcohol** | | | | | | | | | | | | |
| **BL** | 11 (38%) | 4 (14%) | 3 (10%) | 3 (10%) | 4 (14%) | 2 (7%) | 1 (3%) | 0 (0%) | 0 (0%) | 1 (3%) | 0 (0%) | 0 (0%) |
| **6wkFU** | 18 (69%) | 5 (19%) | 1 (4%) | 0 (0%) | 0 (0%) | 0 (0%) | 1 (4%) | 0 (0%) | 0 (0%) | 1 (4%) | 0 (0%) | 0 (0%) |
| **12mFU** | 12 (71%) | 3 (18%) | 1 (5.9%) | 1 (5.9%) | 0 (0%) | 0 (0%) | 0 (0%) | 0 (0%) | 0 (0%) | 0 (0%) | 0 (0%) | 0 (0%) |
| **Caffeine** | | | | | | | | | | | | |
| **BL** | 8 (28%) | 1 (3%) | 1 (3%) | 1 (3%) | 3 (10%) | 1 (3%) | 0 (0%) | 11 (38%) | 3 (10%) | 0 (0%) | 0 (0%) | 0 (0%) |
| **6wkFU** | 17 (65%) | 3 (12%) | 0 (0%) | 1 (4%) | 2 (8%) | 2 (8%) | 0 (0%) | 1 (4%) | 0 (0%) | 0 (0%) | 0 (0%) | 0 (0%) |
| **12mFU** | 6 (35%) | 0 (0%) | 4 (24%) | 1 (5.9%) | 1 (5.9%) | 2 (12%) | 2 (12%) | 1 (5.9%) | 0 (0%) | 0 (0%) | 0 (0%) | 0 (0%) |
| **Tobacco** | | | | | | | | | | | | |

| | | | | | | | | | | | | |
|---|---|---|---|---|---|---|---|---|---|---|---|---|
| **BL** | 29 (100%) | 0 (0%) | 0 (0%) | 0 (0%) | 0 (0%) | 0 (0%) | 0 (0%) | 0 (0%) | 0 (0%) | 0 (0%) | 0 (0%) | 0 (0%) |
| **6wkFU** | 26 (100%) | 0 (0%) | 0 (0%) | 0 (0%) | 0 (0%) | 0 (0%) | 0 (0%) | 0 (0%) | 0 (0%) | 0 (0%) | 0 (0%) | 0 (0%) |
| **12mFU** | 17 | 0 (0%) | 0 (0%) | 0 (0%) | 0 (0%) | 0 (0%) | 0 (0%) | 0 (0%) | 0 (0%) | 0 (0%) | 0 (0%) | 0 (0%) |

Screener reports on previous 30 days. N (%); number (percentage) of participants who selected a given serving size at BL (baseline; N=29), 6wkFU (six-week follow-up; N=26), and 12mFU (12-month follow-up; N=17) visits. The mean (SD) total score was 12 (10), 6 (3), and 6 (4) at BL, 6wkFU, and 12mFU, respectively with scores ranging from zero (100% adherence) to 82 (0% adherence).[2]

#### ST5. Adverse Events Occurring in ≥ 10% of Participants by Treatment Stage and Grade

| | | | Prefeeding | | | Fasting | | | Refeeding | | |
|---|---|---|---|---|---|---|---|---|---|---|---|
| **AE Classification** | **N (%)** | **Total AEs** | **G1** | **G2** | **G3** | **G1** | **G2** | **G3** | **G1** | **G2** | **G3** |
| Fatigue | 26 (90%) | 79 | 2 | 0 | 0 | 46 | 14 | 0 | 12 | 5 | 0 |
| Blood bicarbonate decreased[†] | 21 (72%) | 22 | 0 | 0 | 0 | 22 | 0 | 0 | 0 | 0 | 0 |
| BUN/creatinine decreased[*] | 18 (62%) | 18 | 1 | 0 | 0 | 10 | 0 | 0 | 7 | 0 | 0 |
| Hypertension | 16 (55%) | 27 | 0 | 1 | 0 | 4 | 14 | 7 | 0 | 1 | 0 |
| Nausea | 14 (48%) | 15 | 0 | 0 | 0 | 11 | 2 | 0 | 2 | 0 | 0 |
| Chloride Decreased[*†] | 13 (45%) | 13 | 0 | 0 | 0 | 13 | 0 | 0 | 0 | 0 | 0 |
| Hypoglycemia[†] | 11 (38%) | 12 | 0 | 0 | 0 | 8 | 4 | 0 | 0 | 0 | 0 |
| Creatinine increased | 11 (38%) | 11 | 0 | 0 | 0 | 7 | 0 | 0 | 4 | 0 | 0 |
| Hypokalemia[†] | 11 (38%) | 11 | 0 | 0 | 0 | 11 | 0 | 0 | 0 | 0 | 0 |
| Blood bilirubin increased[†] | 10 (34%) | 10 | 0 | 0 | 0 | 9 | 1 | 0 | 0 | 0 | 0 |
| Dizziness | 10 (34%) | 24 | 0 | 0 | 0 | 22 | 1 | 0 | 1 | 0 | 0 |
| Headache | 9 (31%) | 10 | 1 | 0 | 0 | 7 | 1 | 0 | 1 | 0 | 0 |
| Insomnia | 8 (28%) | 8 | 0 | 0 | 0 | 6 | 1 | 0 | 1 | 0 | 0 |
| BUN Decreased[*] | 8 (28%) | 8 | 0 | 0 | 0 | 3 | 0 | 0 | 5 | 0 | 0 |
| Presyncope[†] | 7 (24%) | 10 | 0 | 0 | 0 | 0 | 10 | 0 | 0 | 0 | 0 |
| AST increased | 6 (21%) | 7 | 0 | 0 | 0 | 5 | 0 | 0 | 2 | 0 | 0 |
| Palpitations | 6 (21%) | 7 | 0 | 0 | 0 | 6 | 0 | 0 | 1 | 0 | 0 |
| ALT increased | 6 (21%) | 6 | 0 | 0 | 0 | 5 | 0 | 0 | 1 | 0 | 0 |
| GERD[†] | 6 (21%) | 6 | 0 | 0 | 0 | 5 | 1 | 0 | 0 | 0 | 0 |
| Hypercalcemia | 6 (21%) | 6 | 0 | 0 | 0 | 5 | 0 | 0 | 1 | 0 | 0 |
| eGFR decreased[*] | 6 (21%) | 6 | 0 | 0 | 0 | 4 | 0 | 0 | 2 | 0 | 0 |
| RBC Increased[*†] | 6 (21%) | 6 | 0 | 0 | 0 | 6 | 0 | 0 | 0 | 0 | 0 |
| Back pain | 5 (17%) | 6 | 0 | 1 | 0 | 3 | 1 | 0 | 1 | 0 | 0 |
| Hemoglobin increased[†] | 5 (17%) | 5 | 0 | 0 | 0 | 5 | 0 | 0 | 0 | 0 | 0 |
| Hyperglycemia | 5 (17%) | 5 | 0 | 0 | 0 | 1 | 0 | 0 | 4 | 0 | 0 |
| Hyponatremia | 4 (14%) | 5 | 0 | 0 | 0 | 4 | 0 | 0 | 1 | 0 | 0 |
| Albumin increased[*] | 4 (14%) | 5 | 0 | 0 | 0 | 2 | 0 | 0 | 3 | 0 | 0 |
| Hematuria | 4 (14%) | 4 | 0 | 0 | 0 | 3 | 0 | 0 | 1 | 0 | 0 |
| Myalgia | 4 (14%) | 4 | 0 | 0 | 0 | 2 | 0 | 0 | 2 | 0 | 0 |
| White blood cell decreased | 4 (14%) | 4 | 0 | 0 | 0 | 1 | 0 | 0 | 2 | 1 | 0 |
| Allergic rhinitis | 4 (14%) | 4 | 1 | 0 | 0 | 3 | 0 | 0 | 0 | 0 | 0 |
| Diarrhea | 3 (10%) | 4 | 0 | 0 | 0 | 0 | 0 | 0 | 4 | 0 | 0 |
| Neutrophil count decreased | 3 (10%) | 4 | 1 | 0 | 0 | 0 | 0 | 0 | 2 | 0 | 1 |
| Abdominal pain[†] | 3 (10%) | 4 | 0 | 0 | 0 | 3 | 1 | 0 | 0 | 0 | 0 |
| Arthralgia[†] | 3 (10%) | 3 | 0 | 0 | 0 | 2 | 1 | 0 | 0 | 0 | 0 |
| Dry mouth[†] | 3 (10%) | 3 | 0 | 0 | 0 | 3 | 0 | 0 | 0 | 0 | 0 |
| Rash maculopapular | 3 (10%) | 3 | 0 | 0 | 0 | 1 | 1 | 0 | 1 | 0 | 0 |

| | | | | | | | | | | | |
|---|---|---|---|---|---|---|---|---|---|---|---|
| Ventricular arrhythmia[†] | 3 (10%) | 3 | 0 | 0 | 0 | 3 | 0 | 0 | 0 | 0 | 0 |
| Hematocrit Increased[*†] | 3 (10%) | 3 | 0 | 0 | 0 | 3 | 0 | 0 | 0 | 0 | 0 |
| A/G Ratio Increased[*] | 3 (10%) | 3 | 0 | 0 | 0 | 2 | 0 | 0 | 1 | 0 | 0 |
| Flatulence[†] | 3 (10%) | 3 | 0 | 0 | 0 | 3 | 0 | 0 | 0 | 0 | 0 |

AE, adverse event; N (%), number (percent) of total participants experiencing AE; Grade 1 (G1), mild; Grade 2 (G2), moderate; Grade 3 (G3), severe; Grade 4 (G4), life-threatening; Grade 5 (G5), death; BUN , blood urea nitrogen; AST, aspartate aminotransferase; ALT, alanine aminotransferase; GERD, gastroesophageal reflux disease; eGFR, estimated glomerular filtration rate; RBC, red blood cell; A/G, albumin/globulin. There were no G4 or G5 events. [*]AE terms are listed in the CTCAE as "other" and were specified and graded according to CTCAE guidelines. [†]Fifteen AEs occurred only during fasting.

#### ST6. Total Adverse Events by Classification, Grade, and Outcome

| | | | | Grade | | | Outcome | | |
|---|---|---|---|---|---|---|---|---|---|
| **SOC** | **AE Term** | **N** | **Total** | **1** | **2** | **3** | **Recovered** | **Persisted** | **Unknown** |
| General disorders and administration site conditions | Fatigue | 26 | 79 | 60 | 19 | | 77 | 2 | |
| Vascular disorders | Hypertension | 16 | 27 | 4 | 16 | 7 | 27 | | |
| Nervous system disorders | Dizziness | 10 | 24 | 23 | 1 | | 23 | 1 | |
| Investigations | Blood bicarbonate decreased | 21 | 22 | 22 | | | 22 | | |
| Metabolism and nutrition disorders | BUN/creatinine decreased[*] | 18 | 18 | 18 | | | 12 | 3 | 3 |
| Gastrointestinal disorders | Nausea | 14 | 15 | 13 | 2 | | 14 | 1 | |
| Metabolism and nutrition disorders | Chloride Decreased[*] | 13 | 13 | 13 | | | 12 | | 1 |
| Metabolism and nutrition disorders | Hypoglycemia | 11 | 12 | 8 | 4 | | 12 | | |
| Investigations | Creatinine increased | 11 | 11 | 11 | | | 9 | 1 | 1 |
| Metabolism and nutrition disorders | Hypokalemia | 11 | 11 | 11 | | | 11 | | |
| Investigations | Blood bilirubin increased | 10 | 10 | 9 | 1 | | 10 | | |
| Nervous system disorders | Headache | 9 | 10 | 9 | 1 | | 9 | 1 | |
| Nervous system disorders | Presyncope | 7 | 10 | | 10 | | 10 | | |
| Psychiatric disorders | Insomnia | 8 | 8 | 7 | 1 | | 7 | 1 | |
| Metabolism and nutrition disorders | BUN Decreased[*] | 8 | 8 | 8 | | | 5 | 2 | 1 |
| Investigations | Aspartate aminotransferase increased | 6 | 7 | 7 | | | 7 | | |
| Cardiac disorders | Palpitations | 6 | 7 | 7 | | | 7 | | |

| | | | | | | | | | |
|---|---|---|---|---|---|---|---|---|---|
| Investigations | Alanine aminotransferase increased | 6 | 6 | 6 | | | 4 | 2 | |
| Gastrointestinal disorders | Gastroesophageal reflux disease | 6 | 6 | 5 | 1 | | 5 | 1 | |
| Metabolism and nutrition disorders | Hypercalcemia | 6 | 6 | 6 | | | 4 | 2 | |
| Investigations | eGFR decreased* | 6 | 6 | 6 | | | 5 | 1 | |
| Investigations | RBC Increased* | 6 | 6 | 6 | | | 6 | | |
| Musculoskeletal and connective tissue disorders | Back pain | 5 | 6 | 4 | 2 | | 6 | | |
| Investigations | Hemoglobin increased | 5 | 5 | 5 | | | 5 | | |
| Metabolism and nutrition disorders | Hyperglycemia | 5 | 5 | 5 | | | 4 | | 1 |
| Metabolism and nutrition disorders | Hyponatremia | 4 | 5 | 5 | | | 5 | | |
| Metabolism and nutrition disorders | Albumin increased* | 4 | 5 | 5 | | | 4 | | 1 |
| Renal and urinary disorders | Hematuria | 4 | 4 | 4 | | | 4 | | |
| Musculoskeletal and connective tissue disorders | Myalgia | 4 | 4 | 4 | | | 4 | | |
| Investigations | White blood cell decreased | 4 | 4 | 3 | 1 | | 3 | | 1 |
| Respiratory, thoracic and mediastinal disorders | Allergic rhinitis | 4 | 4 | 4 | | | 1 | 3 | |
| Gastrointestinal disorders | Diarrhea | 3 | 4 | 4 | | | 2 | 1 | 1 |
| Investigations | Neutrophil count decreased | 3 | 4 | 3 | | 1 | 2 | | 2 |
| Gastrointestinal disorders | Abdominal pain | 3 | 4 | 3 | 1 | | 4 | | |
| Musculoskeletal and connective tissue disorders | Arthralgia | 3 | 3 | 2 | 1 | | 1 | 2 | |
| Gastrointestinal disorders | Dry mouth | 3 | 3 | 3 | | | 3 | | |
| Skin and subcutaneous tissue disorders | Rash maculopapular | 3 | 3 | 2 | 1 | | 3 | | |
| Cardiac disorders | Ventricular arrhythmia | 3 | 3 | 3 | | | 2 | 1 | |
| Investigations | Hematocrit Increased* | 3 | 3 | 3 | | | 3 | | |
| Metabolism and nutrition disorders | A/G Ratio Increased* | 3 | 3 | 3 | | | 3 | | |

| | | | | | | | | | |
|---|---|---|---|---|---|---|---|---|---|
| Gastrointestinal disorders | Flatulence | 3 | 3 | 3 | | | 3 | | |
| Nervous system disorders | Extrapyramidal disorder | 2 | 2 | 2 | | | 2 | | |
| Metabolism and nutrition disorders | Hyperkalemia | 2 | 2 | 2 | | | 1 | 1 | |
| Metabolism and nutrition disorders | Hypophosphatemia | 2 | 2 | 2 | | | | | 2 |
| Musculoskeletal and connective tissue disorders | Neck pain | 2 | 2 | 2 | | | 1 | 1 | |
| Gastrointestinal disorders | Toothache | 2 | 2 | 2 | | | | 2 | |
| Renal and urinary disorders | Urinary tract pain | 2 | 2 | 2 | | | 2 | | |
| Infections and infestations | Vaginal infection | 2 | 2 | 2 | | | 2 | | |
| Gastrointestinal disorders | Vomiting | 2 | 2 | 2 | | | 2 | | |
| Investigations | MCHC increased* | 2 | 2 | 2 | | | 2 | | |
| Investigations | Electrocardiogram QT corrected interval prolonged | 2 | 2 | 2 | | | | 1 | 1 |
| Respiratory, thoracic and mediastinal disorders | Nasal congestion | 1 | 2 | 2 | | | 1 | | 1 |
| Investigations | Alkaline phosphatase increased | 1 | 1 | 1 | | | 1 | | |
| Musculoskeletal and connective tissue disorders | Buttock pain | 1 | 1 | 1 | | | 1 | | |
| Cardiac disorders | Chest pain-cardiac | 1 | 1 | 1 | | | 1 | | |
| Nervous system disorders | Concentration impairment | 1 | 1 | 1 | | | 1 | | |
| Respiratory, thoracic and mediastinal disorders | Cough | 1 | 1 | 1 | | | 1 | | |
| Eye disorders | Dry eye | 1 | 1 | 1 | | | 1 | | |
| Skin and subcutaneous tissue disorders | Dry skin | 1 | 1 | 1 | | | 1 | | |
| Gastrointestinal disorders | Dyspepsia | 1 | 1 | 1 | | | 1 | | |
| Renal and urinary disorders | Dysuria | 1 | 1 | 1 | | | 1 | | |
| General disorders and administration site conditions | Edema face | 1 | 1 | 1 | | | 1 | | |

| | | | | | | | | | |
|---|---|---|---|---|---|---|---|---|---|
| Eye disorders | Hordeolum* | 1 | 1 | 1 | | | 1 | | |
| Gastrointestinal disorders | Gastritis | 1 | 1 | 1 | | | 1 | | |
| Metabolism and nutrition disorders | Hypermagnesemia | 1 | 1 | 1 | | | | | 1 |
| Metabolism and nutrition disorders | Hyperuricemia | 1 | 1 | 1 | | | | | 1 |
| Musculoskeletal and connective tissue disorders | Muscle weakness lower limb | 1 | 1 | 1 | | | 1 | | |
| Infections and infestations | Papulopustular rash | 1 | 1 | 1 | | | 1 | | |
| Investigations | Platelet count decreased | 1 | 1 | 1 | | | 1 | | |
| Renal and urinary disorders | Proteinuria | 1 | 1 | 1 | | | 1 | | |
| Respiratory, thoracic and mediastinal disorders | Sore throat | 1 | 1 | 1 | | | 1 | | |
| Cardiac disorders | Tachycardia | 1 | 1 | 1 | | | 1 | | |
| Renal and urinary disorders | Urinary frequency | 1 | 1 | 1 | | | | 1 | |
| Infections and infestations | Urinary tract infection | 1 | 1 | 1 | | | | 1 | |
| Renal and urinary disorders | Urine discoloration | 1 | 1 | 1 | | | 1 | | |
| Reproductive system and breast disorders | Vaginal inflammation | 1 | 1 | 1 | | | 1 | | |
| Ear and labyrinth disorders | Vertigo | 1 | 1 | | 1 | | 1 | | |
| Eye disorders | Vision decreased | 1 | 1 | | 1 | | 1 | | |
| Investigations | Lymphs Absolute Increased* | 1 | 1 | 1 | | | 1 | | |
| Investigations | RDW Decreased* | 1 | 1 | 1 | | | 1 | | |
| Investigations | RDW Increased* | 1 | 1 | 1 | | | 1 | | |
| Metabolism and nutrition disorders | Anion Gap Increased* | 1 | 1 | 1 | | | | | 1 |
| Metabolism and nutrition disorders | Creatinine decreased* | 1 | 1 | 1 | | | | 1 | |
| Metabolism and nutrition disorders | Insulin decreased* | 1 | 1 | 1 | | | 1 | | |
| Renal and urinary disorders | Urine Analysis Positive leukocytes* | 1 | 1 | 1 | | | | | 1 |

| | | | | | | | | | |
|---|---|---|---|---|---|---|---|---|---|
| Renal and urinary disorders | Urinary hesitancy* | 1 | 1 | 1 | | | 1 | | |
| | **Total** | **359** | **453** | **381** | **64** | **8** | **400** | **33** | **20** |

N, total number of participants experiencing AE; SOC, System Organ Class; AE, adverse event; Grade 1, mild; Grade 2, moderate; Grade 3, severe; Grade 4, life-threatening; Grade 5, death. BUN, blood urea nitrogen; eGFR, estimated glomerular filtration rate; RBC, red blood cell; A/G, albumin/globulin; MCHC, mean corpuscular hemoglobin concentration; RDW, red blood cell distribution width. AEs described as persisted were all G1 events that had not resolved at EOR or 6wkFU. Of the 20 AEs with unknown outcome, 19 were G1 and 1 was G3. There were no Grade 4 or 5 events. *AE terms are listed in the CTCAE as "other" and were specified and graded according to CTCAE guidelines.

#### ST7. CBC by Visit

| | Median (IQR) | | | | |
|---|---|---|---|---|---|
| | **BL** | **EOF** | **EOR** | **FU** | **12mFU** |
| **Hematocrit, %** | | | | | |
| All | 42.4 (39.2, 45.3) | 45.0 (42.9, 47.2) | 43.1 (40.3,45.6) | 41.4 (40.2, 44.1) | 43.3 (38.9, 46.0) |
| 34.0-46.6 (female) | 40.7 (38.2, 42.5) | 43.4 (42.0, 45.0) | 42.2 (38.5, 43.5) | 40.4 (39.0, 41.0) | 40.5 (38.9, 43.4) |
| 37.5-51.0 (male) | 46.8 (44.6, 49.7) | 48.7 (46.5, 51.4) | 46.4 (44.6, 48.4) | 44.3 (42.8, 48.1) | 47.7 (46.8, 49.0) |
| **Hemoglobin, g/dL** | | | | | |
| All | 14.4 (12.8, 15.0) | 15.6 (14.6, 16.6) | 15.0 (13.5, 15.6) | 13.9 (13.1, 15.0) | 14.1 (13.0, 15.3) |
| 11.1-15.9 (female) | 14.2 (12.5, 14.4) | 14.9 (14.1, 15.6) | 14.3 (13.2, 15.4) | 13.4 (12.6, 13.9) | 13.4 (12.9, 14.2) |
| 13.0-17.7 (male) | 16.0 (14.6, 16.9) | 17.0 (16.4, 17.8) | 15.6 (15.1, 16.6) | 15.3 (14.2, 16.4) | 16.4 (15.8, 16.6) |
| **MCV, fL** | | | | | |
| 79-97 | 89 (87, 92) | 88 (85, 90) | 89 (86, 91) | 92 (89, 95) | 92 (90, 95) |
| **MCH, pg** | | | | | |
| 26.6-33.0 | 30.0 (29.2, 31.3) | 30.9 (29.3, 31.5) | 30.6 (29.7, 31.4) | 30.8 (29.6, 31.4) | 30.7 (29.6, 31.3) |
| **MCHC, g/dL** | | | | | |
| 31.5-35.7 | 33.9 (33.1, 34.4) | 34.5 (34.0, 35.1) | 34.3 (33.5, 34.6) | 33.3 (32.7, 34.0) | 33.3 (32.6, 33.7) |
| **RDW, %** | | | | | |
| All | 12.6 (12.1, 13.3) | 12.8 (12.1, 13.4) | 12.8 (12.3, 13.5) | 13.1 (12.6, 13.8) | 12.7 (12.3, 13.3) |
| 11.7-15.4 (female) | 12.6 (12.2, 13.1) | 12.8 (12.3, 13.6) | 12.8 (12.6, 13.5) | 13.2 (12.7, 13.8) | 12.7 (12.3, 13.2)) |
| 11.6-15.4 (male) | 12.4 (11.9, 13.2) | 12.4 (12.0, 13.2) | 12.6 (11.9, 13.3) | 13.0 (12.6, 13.4) | 13.2 (12.6, 13.3) |
| **Platelets, x10$^3$/µL** | | | | | |
| 150-450 | 254 (223, 285) | 240 (204, 304) | 222 (204, 292) | 235 (219, 285) | 266 (220, 307) |
| **RBC count, x10$^6$/µL** | | | | | |
| All | 4.74 (4.43, 5.07) | 5.04 (4.87, 5.39) | 4.89 (4.50, 5.13) | 4.55 (4.26, 4.87) | 4.62 (4.24, 4.99) |
| 3.77-5.28 (female) | 4.48 (4.27, 4.78) | 4.92 (4.74, 5.04) | 4.77 (4.30, 5.00) | 4.44 (4.12, 4.66) | 4.41 (4.18, 4.63) |
| 4.14-5.80 (male) | 5.23 (4.96, 5.66) | 5.48 (5.33, 5.82) | 5.29 (5.01, 5.51) | 4.88 (4.78, 5.06) | 5.40 (4.99, 5.42) |
| **WBC, x10$^3$/µL** | | | | | |
| 3.4-10.8 | 5.4 (4.6, 6.3) | 4.7 (4.2, 5.8) | 4.5 (3.6, 5.4) | 4.8 (4.4, 5.8) | 5.4 (4.6, 6.9) |
| **Neutrophils, %** | 56 (50, 61) | 54 (48, 59) | 48 (43, 56) | 54 (50, 60) | 57 (54, 65) |
| **Neutrophils, x10$^3$/µL** | | | | | |
| 1.4-7.0 | 2.9 (2.5, 3.6) | 2.6 (2.0, 3.1) | 2.1 (1.5, 3.0) | 2.8 (2.3, 3.2) | 3.0 (2.6, 4.2) |
| **Lymphocytes, %** | 31 (27, 38) | 34 (28, 42) | 38 (29, 45) | 33 (28, 38) | 32 (24, 34) |
| **Lymphocytes, x10$^3$/µL** | | | | | |

| | | | | | |
|---|---|---|---|---|---|
| 0.7-3.1 | 1.7 (1.2, 2.1) | 1.6 (1.3, 1.9) | 1.4 (1.3, 1.9) | 1.5 (1.3, 2.1) | 1.5 (1.3, 2.2) |
| **Monocytes, %** | 8 (7, 10) | 10 (8, 11) | 10 (9, 12) | 8 (7, 10) | 8 (7, 9) |
| **Monocytes, x10$^3$/µL**<br>0.1-0.9 | 0.5 (0.4, 0.5) | 0.5 (0.4, 0.5) | 0.4 (0.4, 0.5) | 0.4 (0.3, 0.5) | 0.4 (0.4, 0.5) |
| **Eosinophils, %** | 2 (2, 3) | 2 (1, 3) | 3 (2, 3) | 2 (1, 3) | 2 (1, 3) |
| **Eosinophils, x10$^3$/µL**<br>0.0-0.4 | 0.1 (0.1, 0.2) | 0.1 (0.1, 0.1) | 0.1 (0.1, 0.2) | 0.1 (0.1, 0.2) | 0.2 (0.1, 0.2) |
| **Basophils, %** | 1 (1, 1) | 1 (1, 1) | 1 (1, 1) | 1 (1, 1) | 1 (1, 1) |
| **Basophils, x10$^3$/µL**<br>0.0-0.2 | 0.0 (0.0, 0.1) | 0.0 (0.0, 0.1) | 0.00 (0.0, 0.0) | 0.0 (0.0, 0.0) | 0.0 (0.0, 0.1) |
| **Immature Grans, %** | 0 (0, 0) | 0 (0, 0) | 0 (0, 0) | 0 (0, 0) | 0 (0, 0) |
| **Immature Grans, x10$^3$/µL**<br>0.0-0.1 | 0 (0, 0) | 0 (0, 0) | 0 (0, 0) | 0 (0, 0) | 0.0 (0.0, 0.0) |

Normal reference ranges are listed below the respective variable.[3] At the BL, EOF, EOR visits, there were 29 participants and at the 6wkFU and 12mFU visits there were 26 and 17 participants, respectively. Two participants began refeeding before the EOF blood draw and were excluded from EOF analysis. Due to laboratory errors, one value for platelets was missing from the BL, EOR, and 6wkFU analysis and 2 values for immature grans were missing from EOR analysis. CBC, complete blood count; IQR, interquartile range; g/dL, grams per deciliter; MCV, Mean corpuscular volume; fL, femtoliter; MCH, mean corpuscular hemoglobin; pg, picogram; MCHC, Mean corpuscular hemoglobin concentration; RDW, red cell distribution width; RBC, red blood cell; µL, microliter; WBC, white blood count; Grans, granulocytes; BL, baseline; EOF, end-of-fast; EOR; end-of-refeed; 6wkFU, six-week follow-up; 12mFU, 12-month follow-up.

#### ST8. Significance of Difference for CBC

| | Estimates (95% CI) | | | | | | |
|---|---|---|---|---|---|---|---|
| | **EOF - BL** | **EOR - BL** | **6wkFU - BL** | **12mFU - BL** | **12mFU – 6wkFU** | **EOR – EOF** | **6wkFU – EOR** |
| **ξ Hematocrit, %** | 2.62*<br>(1.80, 3.44) | 0.35<br>(-0.46, 1.17) | -0.32<br>(-1.17, 0.53) | 0.79<br>(-0.19, 1.77) | 1.11*<br>(0.11, 2.10) | -2.26*<br>(-3.08, -1.44) | -0.67<br>(-1.52, 0.18) |
| **Hemoglobin, g/dL** | 1.26*<br>(0.95, 1.57) | 0.29<br>(-0.03, 0.60) | -0.30<br>(-0.63, 0.02) | 0.06<br>(-0.31, 0.44) | 0.36<br>(-0.02, 0.74) | -0.97*<br>(-1.29, -0.66) | -0.59*<br>(-0.91, -0.26) |
| **MCV, fL** | -1.24*<br>(-2.02, -0.47) | -0.41<br>(-1.19, 0.36) | 2.57*<br>(1.76, 3.37) | 2.70*<br>(1.77, 3.63) | 0.13<br>(-0.81, 1.08) | 0.83*<br>(0.05, 1.60) | 2.98*<br>(2.18, 3.79) |
| **MCH, pg** | 0.27*<br>(0.01, 0.54) | 0.20<br>(-0.07, 0.46) | 0.39*<br>(0.12, 0.66) | 0.49*<br>(0.17, 0.81) | 0.10<br>(-0.22, 0.42) | -0.08<br>(-0.34, 0.19) | 0.19<br>(-0.08, 0.46) |
| **MCHC, g/dL** | 0.75*<br>(0.46, 1.04) | 0.33*<br>(0.04, 0.62) | -0.54*<br>(-0.84, -0.24) | -0.46*<br>(-0.81, -0.11) | 0.08<br>(-0.28, 0.43) | -0.41*<br>(-0.70, -0.12) | -0.87*<br>(-1.17, -0.57) |
| **ξ RDW, %** | 0.27*<br>(0.07, 0.46) | 0.26*<br>(0.07, 0.46) | 0.49*<br>(0.29, 0.70) | 0.07<br>(-0.16, 0.31) | -0.42*<br>(-0.66, -0.18) | -0.01<br>(-0.20, 0.19) | 0.23*<br>(0.03, 0.43) |
| **Platelets, x103/µL** | 5.45<br>(-8.01, 18.78) | -18.46*<br>(-31.88, -5.05) | -12.61<br>(-26.55, 1.32) | -9.19<br>(-25.02, 6.78) | 3.42<br>(-12.64, 19.63) | -23.92*<br>(-37.25, -10.46) | 5.86<br>(-8.08, 19.78) |
| **RBC count, x10$^6$/µL** | 0.38*<br>(0.27, 0.49) | 0.07<br>(-0.04, 0.18) | -0.16*<br>(-0.28, -0.05) | -0.06<br>(-0.19, 0.07) | 0.10<br>(-0.03, 0.24) | -0.31*<br>(-0.42, -0.20) | -0.23*<br>(-0.34, -0.11) |
| **WBC, x103/µL** | -0.28<br>(-0.65, 0.08) | -0.79*<br>(-1.16, -0.43) | -0.32<br>(-0.69, 0.06) | -0.00<br>(-0.43, 0.44) | 0.32<br>(-0.12, 0.76) | -0.51*<br>(-0.88, -0.15) | 0.48*<br>(0.10, 0.86) |
| **Neutrophils, %** | -1.72<br>(-4.28, 0.83) | -5.97*<br>(-8.52, -3.41) | -0.73<br>(-3.37, 1.94) | 2.22<br>(-0.83, 5.30) | 2.95<br>(-0.16, 6.06) | -4.24*<br>(-6.80, -1.69) | 5.24*<br>(2.60, 7.90) |
| **ξ Neutrophils, x10$^3$/µL** | -0.27*<br>(-0.53, -0.01) | -0.76*<br>(-1.02, -0.50) | -0.23<br>(-0.50, 0.04) | 0.02<br>(-0.29, 0.34) | 0.25<br>(-0.07, 0.57) | -0.49*<br>(-0.75, -0.23) | 0.53*<br>(0.26, 0.80) |
| **Lymphocytes, %** | 1.45<br>(-0.73, 3.63) | 4.31*<br>(2.13, 6.49) | 0.39<br>(-1.88, 2.64) | -1.77<br>(-4.40, 0.84) | -2.16<br>(-4.82, 0.49) | 2.86*<br>(0.68, 5.04) | -3.92*<br>(-6.19, -1.67) |
| **Lymphocytes, x103/µL** | 0.00<br>(-0.13, 0.13) | -0.08<br>(-0.21, 0.05) | -0.08<br>(-0.21, 0.06) | -0.15<br>(-0.30, 0.01) | -0.07<br>(-0.23, 0.09) | -0.08<br>(-0.21, 0.05) | 0.00<br>(-0.13, 0.14) |
| **λMonocytes, %** | 1.08<br>(0.91, 1.27) | 1.16<br>(0.98, 1.36) | 1.00<br>(0.84, 1.20) | 0.92<br>(0.75, 1.13) | 0.92<br>(0.74, 1.13) | 1.07<br>(0.91, 1.26) | 0.87<br>(0.73, 1.03) |
| **λMonocytesτ, x103/µL** | 1.05<br>(0.83, 1.34) | 1.02<br>(0.81, 1.30) | 0.97<br>(0.75, 1.24) | 0.97<br>(0.73, 1.29) | 1.00<br>(0.75, 1.34) | 0.97<br>(0.77, 1.23) | 0.95<br>(0.74, 1.21) |
| **λEosinophils, %** | 0.89<br>(0.63, 1.24) | 1.13<br>(0.82, 1.55) | 1.07<br>(0.76, 1.50) | 1.09<br>(0.74, 1.60) | 1.02<br>(0.69, 1.50) | 1.27<br>(0.92, 1.76) | 0.95<br>(0.68, 1.32) |

| | | | | | | | |
|---|---|---|---|---|---|---|---|
| **[λ]Eosinophils[τ], x103/µL** | 0.81<br>(0.50, 1.31) | 0.97<br>(0.62, 1.54) | 1.07<br>(0.67, 1.69) | 1.23<br>(0.74, 2.05) | 1.15<br>(0.69, 1.92) | 1.20<br>(0.74, 1.94) | 1.10<br>(0.69, 1.75) |
| **[λ]Basophils, %** | 0.93<br>(0.54, 1.60) | 0.96<br>(0.56, 1.65) | 0.95<br>(0.55, 1.66) | 0.94<br>(0.50, 1.77) | 0.99<br>(0.52, 1.90) | 1.04<br>(0.60, 1.80) | 0.99<br>(0.56, 1.73) |
| **[‡]Basophils, $x10^3$/µL** | 1.92<br>(0.38, 9.67) | 0.69<br>(0.13, 3.72) | 0.58<br>(0.10, 3.41) | 6.12<br>(0.79, 47.8) | 10.50[α]<br>(1.15, 96.90) | 0.34<br>(0.06, 1.85) | 0.79<br>(0.13, 4.87) |
| **Immature Grans, %** | NR | NR | NR | NR | NR | NR | NR |
| **Immature Grans, x103/µL** | NR | NR | NR | NR | NR | NR | NR |

CBC, complete blood count; CI, confidence interval; BL, baseline; EOF, end-of-fast; EOR, end-of-refeed; 6wkFU, six-week follow-up; 12mFU, 12-month follow-up; g/dL, grams per deciliter; MCV, Mean Corpuscular Volume; fL, femtoliter; MCH, mean corpuscular hemoglobin; pg, picogram; MCHC, Mean corpuscular hemoglobin concentration; RDW, red cell distribution width; RBC, red blood cell; µL, microliter; WBC, white blood count; Grans, granulocytes; NR, Results not reported due to poor diagnostics. [*]Zero lies outside the 95% CI so the finding is considered significant. [α]One lies outside the 95% CI so the finding is considered statistically significant. [‡]Logistic regression (included baseline medication-controlled SBP/DBP indicator as a control variable); [λ]Poisson regression; [τ]Measure was multiplied by 10 prior to statistical modeling; [ξ]Used robust mixed-effects model on complete cases.

#### ST9. CMP by Visit

| | Median (IQR) | | | | |
|---|---|---|---|---|---|
| | **BL** | **EOF** | **EOR** | **FU** | **12mFU** |
| **ALT, IU/L** | | | | | |
| All | 19 (15, 26) | 22 (17, 32) | 26 (20, 36) | 18 (14, 26) | 18 (15, 21) |
| 0-32 (female) | 17 (14, 20) | 24 (17, 30) | 26 (19, 37) | 17 (14, 24) | 18 (14, 21) |
| 0-44 (male) | 26 (20, 37) | 22 (18, 31) | 24 (20, 28) | 21 (18, 26) | 18 (18, 19) |
| **AST, IU/L** | | | | | |
| 0-40 | 22 (18, 27) | 30 (25, 36) | 32 (24, 38) | 21 (18, 23) | 22 (18, 23) |
| **Albumin, g/L** | | | | | |
| All | 46 (44, 48) | 47 (45, 49) | 45 (43, 47) | 46 (42, 46) | 44 (42, 47) |
| 38-48 (31-50 y, female) | 42 (42, 42) | 44 (44, 44) | 42 (42, 42) | 45 (45, 45) | 44 (N/A) |
| 40-50 (31-50 y, male) | 50 (50, 50) | 50 (50, 50) | 47 (47, 47) | N/A | N/A |
| 38-49 (51-60 y) | 47 (46, 49) | 48 (45, 50) | 48 (44, 49) | 44 (41, 46) | 44 (43, 47) |
| 38-48 (61-70 y) | 45 (44, 47) | 47 (45, 48) | 44 (43, 46) | 46 (44, 46) | 45 (42, 47) |
| 37-47 (71-80 y) | N/A | N/A | N/A | N/A | 43 (42, 44) |
| **Globulin, g/dL** | | | | | |
| 1.5-4.5 | 2.5 (2.2, 2.7) | 2.5 (2.3, 2.9) | 2.4 (2.2, 2.7) | 2.3 (2.1, 2.6) | 2.5 (2.3, 2.8) |
| **A/G Ratio** | | | | | |
| 1.2-2.2 | 1.9 (1.7, 2.1) | 1.9 (1.7, 2.1) | 1.9 (1.7, 2.2) | 1.9 (1.8, 2.1) | 1.9 (1.5, 2.0) |
| **Alkaline Phosphate, IU/L** | | | | | |
| 44-121 | 87 (70, 96) | 90 (70, 104) | 82 (67, 94) | 91 (84, 101) | 91 (71, 106) |
| **Bilirubin, µmol/L** | | | | | |
| 0.0-20.5 | 10.3 (6.8, 13.7) | 15.4 (10.3, 21.4) | 6.8 (5.1, 8.6) | 8.6 (6.8, 10.3) | 6.8 (5.1, 8.6) |
| **BUN, mg/dL** | | | | | |
| All | 9 (8, 10) | 8 (7, 10) | 6 (5, 8) | 8 (7, 10) | 14 (13, 17) |
| 6-24 (40-59 y) | 9 (8, 10) | 8 (7, 10) | 7 (6, 7) | 10 (8, 10) | 13 (10, 16) |
| 8-27 (60-89 y) | 9 (8, 10) | 8 (7, 10) | 6 (4, 8) | 8 (7, 10) | 10 (9, 13) |
| **Creatinine, mg/dL** | | | | | |
| All | 0.80 (0.77, 0.92) | 1.00 (0.84, 1.14) | 0.92 (0.77, 1.06) | 0.77 (0.67, 0.90) | 0.78 (0.64, 0.90) |
| 0.57-1.00 (female) | 0.80 (0.72, 0.82) | 0.91 (0.77, 1.00) | 0.84 (0.75, 0.99) | 0.72 (0.65, 0.79) | 0.75 (0.63, 0.81) |
| 0.76-1.27 (male) | 0.99 (0.81, 1.10) | 1.14 (1.04, 1.19) | 1.07 (0.94, 1.30) | 0.91 (0.84, 1.04) | 0.95 (0.88, 0.98) |
| **BUN/Creatinine Ratio** | | | | | |
| All | 11 (9, 13) | 8 (7, 11) | 7 (6, 9) | 11 (9, 13) | 14 (13, 17) |

| | | | | | |
|---|---|---|---|---|---|
| 9-23 (18-59 y, female) | 10 (8, 15) | 8 (7, 8) | 8 (7, 10) | 12 (9, 15) | 16 (13, 20) |
| 12-28 (>59 y, female) | 11 (9, 13) | 8 (7, 11) | 6 (5, 7) | 12 (9, 13) | 16 (13, 17) |
| 9-20 (18-59 y, male) | 10 (9, 11) | 8 (8, 9) | 4 (4, 6) | 9 (8, 10) | N/A |
| 10-24 (>59 y, male) | 12 (11, 14) | 10 (9, 12) | 8 (8, 10) | 11 (10, 11) | 13 (10, 14) |
| **Calcium, mmol/L** | | | | | |
| All | 2.40 (2.33, 2.45) | 2.50 (2.42, 2.55) | 2.45 (2.40, 2.50) | 2.39 (2.29, 2.49) | 2.38 (2.34, 2.43) |
| 2.18-2.55 (18-59 y) | 2.39 (2.34, 2.52) | 2.48 (2.42, 2.55) | 2.46 (2.41, 2.49) | 2.39 (2.32, 2.41) | 2.35 (2.29, 2.41) |
| 2.18-2.56 (>59 y, female) | 2.42 (2.35, 2.45) | 2.54 (2.45, 2.58) | 2.48 (2.40, 2.50) | 2.42 (2.37, 2.49) | 2.38 (2.36, 2.41) |
| 2.15-2.55 (>59 y, male) | 2.35 (2.31, 2.39) | 2.48 (2.44, 2.51) | 2.42 (2.38, 2.45) | 2.34 (2.31, 2.37) | 2.38 (2.35, 2.50) |
| **Carbon Dioxide, mmol/L** | | | | | |
| 20-29 | 24 (21, 26) | 17 (15, 20) | 25 (23, 26) | 24 (22, 26) | 24 (23, 24) |
| **eGFR, mL/min/1.73** | | | | | |
| >59 | 82 (75, 93) | 71 (63, 78) | 75 (62, 86) | 90 (79, 95) | 89 (85, 98) |
| **Glucose, mg/dL** | | | | | |
| 65-99 | 95 (88, 101) | 74 (69, 80) | 96 (91, 105) | 92 (87, 98) | 91 (84, 96) |
| **Protein, g/dL** | | | | | |
| 6.0-8.5 | 7.0 (6.7, 7.3) | 7.3 (7.0, 7.6) | 6.9 (6.6, 7.2) | 6.7 (6.6, 7.0) | 7.0 (6.7, 7.2) |
| **Potassium, mmol/L** | | | | | |
| 3.5-5.2 | 4.2 (4.1, 4.4) | 3.6 (3.4, 3.9) | 4.2 (4.1, 4.5) | 4.2 (4.1, 4.5) | 4.5 (4.0, 4.7) |
| **Sodium, mmol/L** | | | | | |
| 134-144 | 141 (139, 143) | 138 (136, 140) | 137 (137, 140) | 142 (141, 143) | 140 (139, 142) |
| **Chloride, mmol/L** | | | | | |
| 96-106 | 102 (101, 104) | 96 (94, 98) | 98 (96, 100) | 104 (103, 106) | 103 (102, 104) |

Normal reference ranges are listed below the respective variable.[3] At the BL, EOF, EOR visits, there were 29 participants and at the 6wkFU and 12mFU visits, there were 26 and 17 participants, respectively. Two participants began refeeding before the EOF blood draw and were excluded from EOF analysis. Due to laboratory errors, 1 value was missing for sodium, potassium, chloride, carbon dioxide, calcium, and ALP at the 12mFU visit. CMP, comprehensive metabolic panel; IQR, interquartile range; BL, baseline; EOF, end-of-fast; EOR, end-of-refeed; 6wkFU, six-week follow-up; 12mFU, 12-month follow-up; ALT, alanine aminotransferase; IU/L, international unit per liter; g/L, gram per liter; g/dL, gram per deciliter; N/A, not applicable; y, years; A/G, Albumin/Globulin; AST, aspartate aminotransferase; µmol/L, micromole per liter; BUN, Blood urea nitrogen; mg/dL, milligram per deciliter; mmol/L, millimol per liter; eGFR, estimated glomerular filtration rate; mL/min, milliliter per minute.

#### ST10. Significance of Difference for CMP

| | Estimates (95% CI) | | | | | | |
|---|---|---|---|---|---|---|---|
| | **EOF -BL** | **EOR - BL** | **6wkFU - BL** | **12mFU - BL** | **12mFU – 6wkFU** | **EOR - EOF** | **6wkFU - EOR** |
| **ALT, IU/L‡** | 5.01*<br>(1.06, 8.96) | 5.68*<br>(1.74, 9.63) | -1.36<br>(-5.44, 2.73) | -2.17<br>(-6.88, 2.54) | -0.82<br>(-5.60, 3.96) | 0.67<br>(-3.28, 4.62) | -7.04*<br>(-11.13, -2.95) |
| **AST, IU/L‡** | 8.34*<br>(5.41, 11.27) | 7.36*<br>(4.43, 10.29) | -1.33<br>(-4.37, 1.70) | -1.36<br>(-4.94, 2.22) | -0.02<br>(-3.65, 3.60) | -0.98<br>(-3.91, 1.95) | -8.69*<br>(-11.73, -5.66) |
| **Albumin, g/L** | 1.24*<br>(0.11, 2.38) | -0.66<br>(-1.79, 0.48) | -1.14<br>(-2.32, 0.03) | -0.76<br>(-2.13, 0.58) | 0.38<br>(-1.01, 1.74) | -1.90*<br>(-3.03, -0.76) | -0.48<br>(-1.66, 0.69) |
| **Globulin, g/dL** | 0.11*<br>(0.01, 0.21) | -0.04<br>(-0.14, 0.05) | -0.10<br>(-0.20, 0.01) | -0.03<br>(-0.15, 0.09) | 0.06<br>(-0.06, 0.19) | -0.16*<br>(-0.25, -0.06) | -0.05<br>(-0.15, 0.05) |
| **A/G Ratio** | -0.04<br>(-0.13, 0.06) | -0.00<br>(-0.10, 0.09) | 0.01<br>(-0.09, 0.11) | 0.00<br>(-0.11, 0.12) | -0.00<br>(-0.12, 0.11) | 0.03<br>(-0.06, 0.13) | 0.01<br>(-0.09, 0.11) |
| **Alkaline Phosphate, IU/L** | 3.24<br>(-0.80, 7.28) | -4.45*<br>(-8.49, -0.41) | 3.15<br>(-1.03, 7.35) | 4.27<br>(-0.69, 9.23) | 1.12<br>(-3.91, 6.14) | -7.69*<br>(-11.73, -3.65) | 7.60*<br>(3.42, 11.79) |
| **Bilirubin, µmol/L‡** | 4.84*<br>(3.59, 6.09) | -2.95*<br>(-4.21, -1.70) | -1.54*<br>(-2.83, -0.24) | -2.34*<br>(-3.84, -0.84) | -0.80<br>(-2.32, 0.72) | -7.79*<br>(-9.04, -6.54) | 1.42*<br>(0.12, 2.71) |
| **BUN, mg/dL‡** | -0.36<br>(-1.37, 0.65) | -2.48*<br>(-3.49, -1.47) | -0.26<br>(-1.30, 0.79) | 2.00*<br>(0.80, 3.20) | 2.26*<br>(1.04, 3.48) | -2.12*<br>(-3.13, -1.11) | 2.22*<br>(1.18, 3.27) |
| **Creatinine, mg/dL** | 0.15*<br>(0.10, 0.20) | 0.10*<br>(0.05, 0.15) | -0.02<br>(-0.07, 0.03) | -0.03<br>(-0.09, 0.02) | -0.01<br>(-0.07, 0.05) | -0.04<br>(-0.09, 0.00) | -0.13*<br>(-0.18, -0.07) |

| | | | | | | | |
|---|---|---|---|---|---|---|---|
| **BUN/Creatinine Ratio** | -2.17*<br>(-3.66, -0.69) | -4.07*<br>(-5.55, -2.59) | 0.34<br>(-1.19, 1.88) | 3.34*<br>(1.60, 5.12) | 3.01*<br>(1.23, 4.80) | -1.90*<br>(-3.38, -0.41) | 4.41*<br>(2.88, 5.94) |
| **Calcium, mmol/L** | 0.10*<br>(0.06, 0.13) | 0.06*<br>(0.03, 0.10) | 0.00<br>(-0.04, 0.04) | -0.04<br>(-0.08, 0.01) | -0.04<br>(-0.08, 0.01) | -0.04*<br>(-0.07, -0.00) | -0.06*<br>(-0.10, -0.02) |
| **Carbon Dioxide, mmol/L‡** | -5.86*<br>(-7.38, -4.35) | 0.85<br>(-0.66, 2.37) | 0.23<br>(-1.33, 1.78) | 0.14<br>(-1.65, 1.94) | -0.08<br>(-1.92, 1.75) | 6.72*<br>(5.20, 8.23) | -0.63<br>(-2.19, 0.93) |
| **eGFR, mL/min/1.73** | -12.07*<br>(-15.96, -8.18) | -7.93*<br>(-11.82, -4.04) | 3.34<br>(-0.69, 7.37) | 6.19*<br>(1.53, 10.84) | 2.84<br>(-1.88, 7.57) | 4.14*<br>(0.25, 8.03) | 11.27*<br>(7.24, 15.30) |
| **Glucose, mg/dL** | -17.10*<br>(-21.63, -12.57) | 3.52<br>(-1.01, 8.05) | -1.38<br>(-6.07, 3.30) | -4.07<br>(-9.44, 1.36) | -2.69<br>(-8.14, 2.82) | 20.62*<br>(16.09, 25.15) | -4.90*<br>(-9.59, -0.21) |
| **Protein, g/dL** | 0.23*<br>(0.09, 0.38) | -0.11<br>(-0.26, 0.04) | -0.21*<br>(-0.36, -0.06) | -0.10<br>(-0.27, 0.08) | 0.11<br>(-0.07, 0.29) | -0.34*<br>(-0.49, -0.20) | -0.10<br>(-0.25, 0.05) |
| **Potassium, mmol/L** | -0.55*<br>(-0.70, -0.39) | 0.03<br>(-0.12, 0.18) | 0.09<br>(-0.07, 0.25) | 0.15<br>(-0.04, 0.34) | 0.06<br>(-0.13, 0.25) | 0.58*<br>(0.43, 0.73) | 0.06<br>(-0.10, 0.22) |
| **Sodium, mmol/L** | -2.79*<br>(-4.06, -1.53) | -2.79*<br>(-4.06, -1.53) | 0.74<br>(-0.57, 2.04) | -0.52<br>(-2.05, 1.00) | -1.26<br>(-2.82, 0.29) | 0.00<br>(-1.26, 1.26) | 3.53*<br>(2.23, 4.83) |
| **Chloride, mmol/L** | -6.10*<br>(-7.39, -4.82) | -4.07*<br>(-5.35, -2.79) | 1.52*<br>(0.20, 2.85) | 0.98<br>(-0.59, 2.51) | -0.54<br>(-2.14, 1.01) | 2.03*<br>(0.75, 3.32) | 5.59*<br>(4.27, 6.92) |

CMP, comprehensive metabolic panel; CI, confidence interval; BL, baseline; EOF, end-of-fast; EOR, end-of-refeed; 6wkFU, SIX-week follow-up; 12mFU, 12-month follow-up; ALT, alanine aminotransferase; IU/L, international unit per liter; g/L, gram per liter; g/dL, gram per deciliter; N/A, not applicable; y, years; A/G, Albumin/Globulin; AST, aspartate aminotransferase; µmol/L, micromole per liter; BUN, Blood urea nitrogen; mg/dL, milligram per deciliter; mmol/L, millimol per liter; eGFR, estimated glomerular filtration rate; mL/min, milliliter per minute. *Zero lies outside the 95% CI so the finding is considered significant. ‡Used robust mixed-effects model on complete cases.

#### ST11. 24-hour Dipstick Urinalysis By Visit

| | | N (%) | | |
|---|---|---|---|---|
| | | **BL** | **EOF** | **EOR** |
| Leukocytes, cells/µL <15 | **0** | 26 (90%) | 24 (83%) | 25 (86%) |
| | **15** | 0 (0%) | 1 (3%) | 0 (0%) |
| | **70** | 1 (3%) | 4 (14%) | 2 (7%) |
| | **125** | 2 (7%) | 0 (0%) | 2 (7%) |
| Nitrite | **Negative** | 24 (83%) | 29 (100%) | 29 (100%) |
| | **Positive** | 5 (17%) | 0 (0%) | 0 (0%) |
| Urobilinogen, µmol/L <17 | **0** | 19 (66%) | 19 (66%) | 20 (69%) |
| | **3.2** | 9 (31%) | 10 (34%) | 9 (31%) |
| | **16** | 1 (3.4%) | 0 (0%) | 0 (0%) |
| Protein, mg/dL <30 | **0** | 27 (93%) | 17 (59%) | 27 (93%) |
| | **15** | 2 (7%) | 9 (31%) | 2 (7%) |
| | **30** | 0 (0%) | 3 (10%) | 0 (0%) |
| pH | **5.0** | 6 (21%) | 25 (86%) | 8 (28%) |
| | **6.0** | 6 (21%) | 4 (14%) | 5 (17%) |
| | **6.5** | 9 (31%) | 0 (0%) | 8 (28%) |
| | **7.0** | 8 (28%) | 0 (0%) | 6 (21%) |
| | **7.5** | 0 (0%) | 0 (0%) | 2 (7%) |
| Blood, cells/µL | **Inconclusive** | 1 (3%)* | 0 (0%) | 0 (0%) |
| | **Negative** | 25 (86%) | 23 (79%) | 26 (90%) |
| | **Positive** | 3 (10%) | 6 (21%) | 3 (10%) |
| Specific Gravity | **1.005** | 6 (21%) | 7 (24%) | 12 (41%) |
| | **1.010** | 16 (55%) | 11 (38%) | 14 (48%) |
| | **1.015** | 7 (24%) | 7 (24%) | 3 (10%) |
| | **1.020** | 0 (0%) | 2 (6.9%) | 0 (0%) |
| | **1.025** | 0 (0%) | 2 (6.9%) | 0 (0%) |
| | **1.030** | 0 (0%) | 2 (6.9%) | 0 (0%) |
| Ketone, mmol/L | **0** | 25 (86%) | 0 (0%) | 27 (93%) |

| | | | | |
|---|---|---|---|---|
| <0.5 | **0.5** | 3 (10%) | 1 (3.4%)[†] | 0 (0%) |
| | **1.5** | 1 (3.4%) | 2 (6.9%) | 1 (3.4%) |
| | **4.0** | 0 (0%) | 3 (10%) | 1 (3.4%) |
| | **8.0** | 0 (0%) | 14 (48%) | 0 (0%) |
| | **16** | 0 (0%) | 9 (31%) | 0 (0%) |
| Bilirubin, µmol/L | **0** | 29 (100%) | 26 (90%) | 29 (100%) |
| <17 | **17** | 0 (0%) | 3 (10%) | 0 (0%) |
| Glucose, mmol/L | **0** | 28 (97%) | 29 (100%) | 28 (97%) |
| <5 | **5** | 1 (3%) | 0 (0%) | 1 (3.4%) |

Normal reference ranges are listed below the respective variable. N (%), number (percent) of participants; BL, baseline; EOF, end-of-fast; EOR, end-of-refeed; µL, microliter; µmol/L, micromole per liter; mg/dL, milligram per deciliter; pH, potential hydrogen; mmol/L, millimol per liter. [*]Participant was menstruating. [†]Participant interrupted majority of the fast with juice.

#### ST12. Daily Rate of Change for Body Weight, Blood Pressure, and Vital Signs

| | Rate of Change/Day ($N_{obs}$, SE, P-value) | | |
|---|---|---|---|
| | **Prefeed** | **Fasting** | **Refeed** |
| **BW, kg** | -1.11<br>(47, 0.18, <0.0001)[*§] | -0.54<br>(342, 0.03, <0.0001)[*] | 0.23<br>(174, 0.03, <0.0001)[*] |
| Early period[‡] | N/A | -0.89<br>(342, 0.03, <0.0001)[*] | N/A |
| Late period[‡] | N/A | -0.43<br>(342, 0.03, <0.0001)[*] | N/A |
| **SBP, mmHg** | -9.04<br>(49, 2.87, 0.005) | -1.21<br>(351, 0.27, <0.0001) | 0.32<br>(184, 0.22, 0.1)[§] |
| **DBP, mmHg** | -1.66<br>(49, 1.28, 0.2)[§] | -0.29<br>(351, 0.13, 0.03) | 0.21<br>(184, 0.12, 0.07)[§] |
| **Pulse, $min^{-1}$** | -1.35<br>(49, 1.25, 0.3)[§] | 0.55<br>(350, 0.16, 0.001) | -0.14<br>(184, 0.19, 0.5)[‡] |
| **$SpO_2$, %** | 0.21<br>(45, 0.22, 0.4)[†§] | 0.01<br>(338, 0.01, 0.3)[§] | -0.01<br>(177, 0.02, 0.6)[§] |
| **BT, °C** | -0.01<br>(47, 0.05, 0.9)[*§] | -0.01<br>(349, 0.00, <0.0001)[§] | 0.01<br>(181, 0.00, 0.02)[§] |

N = 27 participants. $N_{obs}$, Total number of observations in model; SE, Standard error of measurement; P-value, probability value (≤ 0.05 is considered significant); SBP, systolic blood pressure; DBP, diastolic blood pressure; mmHg, milliliter mercury; BW, body weight; kg, kilogram; min-1, per minute; SpO2, saturation of peripheral oxygen; %, percent; BT, body temperature; °C, degrees Celsius; N/A, not applicable. [*]26 participants. [†]24 participants. [‡]Estimated daily rate of change of BW in early period (day 1-5) and late period (>5 days) of fasting. [§]Model excluded random slope.

#### ST13. Significance of Difference for Cardiometabolic Biomarkers

| | Estimates (95% CI) | | | | | | |
|---|---|---|---|---|---|---|---|
| | **EOF - BL** | **EOR - BL** | **6wkFU - BL** | **12mFU - BL** | **12mFU – 6wkFU** | **EOR - EOF** | **6wkFU - EOR** |
| **Weight, kg[‡]** | -7.76[*]<br>(-8.84, -6.67) | -6.59[*]<br>(-7.68, -5.50) | -6.72[*]<br>(-7.87, -5.58) | -5.41[*]<br>(-6.72, -4.11) | 1.31<br>(-0.02, 2.64) | 1.17[*]<br>(0.08, 2.25) | -0.13<br>(-1.28, 1.01) |
| **BMI, kg/m2** | -2.73[*]<br>(-3.41, -2.05) | -2.33[*]<br>(-3.01, -1.65) | -2.58[*]<br>(-3.29, -1.86) | -2.67[*]<br>(-3.49, -1.86) | -0.09<br>(-0.93, 0.74) | 0.40<br>(-0.28, 1.08) | -0.25<br>(-0.97, 0.46) |
| **AC, cm[‡]** | -7.45[*]<br>(-8.83, -6.07) | -5.94[*]<br>(-7.32, -4.56) | -6.55[*]<br>(-8.00, -5.10) | -7.92[*]<br>(-9.58, -6.26) | -1.37<br>(-3.06, 0.33) | 1.51[*]<br>(0.13, 2.89) | -0.61<br>(-2.06, 0.84) |

| | | | | | | | |
|---|---|---|---|---|---|---|---|
| **SBP, mmHg** | -27.90* (-34.81, -20.99) | -27.38* (-34.29, -20.47) | -19.19* (-26.32, -12.03) | -17.20* (-25.42, -8.99) | 1.99 (-6.38, 10.33) | 0.52 (-6.39, 7.43) | 8.19* (1.06, 15.35) |
| **DBP, mmHg** | -7.05* (-10.59, -3.52) | -6.55* (-10.09, -3.02) | -5.13* (-8.77, -1.46) | -8.89* (-13.09, -4.67) | -3.76 (-8.04, 0.51) | 0.50 (-3.03, 4.03) | 1.43 (-2.22, 5.09) |
| **Total Cholesterol, mol/L** | 0.30 (-0.03, 0.63) | -0.34* (-0.67, -0.02) | -0.26 (-0.60, 0.08) | 0.10 (-0.30, 0.48) | 0.35 (-0.05, 0.75) | -0.64* (-0.97, -0.32) | 0.08 (-0.26, 0.42) |
| **LDL, mmol/L** | 0.38* (0.09, 0.67) | -0.42* (-0.71, -0.14) | -0.23 (-0.53, 0.07) | 0.04 (-0.31, 0.38) | 0.27 (-0.09, 0.62) | -0.80* (-1.09, -0.51) | 0.19 (-0.11, 0.49) |
| **HDL, mmol/L** | -0.06 (-0.15, 0.03) | -0.04 (-0.13, 0.05) | -0.03 (-0.12, 0.07) | 0.11* (0.00, 0.22) | 0.14* (0.03, 0.24) | 0.02 (-0.07, 0.10) | 0.02 (-0.08, 0.11) |
| **VLDL, mmol/L** | -0.02 (-0.10, 0.06) | 0.12* (0.04, 0.20) | -0.00 (-0.08, 0.08) | -0.06 (-0.15, 0.03) | -0.05 (-0.15, 0.04) | 0.14* (0.06, 0.22) | -0.13* (-0.21, -0.05) |
| **Triglycerides, mmol/L** | -0.06 (-0.26, 0.13) | 0.33* (0.14, 0.53) | 0.00 (-0.20, 0.20) | -0.16 (-0.39, 0.07) | -0.16 (-0.40, 0.07) | 0.40* (0.20, 0.59) | -0.33* (-0.53, -0.13) |
| **Insulin, pmol/L‡** | -19.19* (-28.72, -9.65) | 11.14* (1.60, 20.68) | 7.12 (-2.76, 17.01) | 2.98 (-8.44, 14.39) | -4.15 (-15.73, 7.43) | 30.33* (20.79, 39.87) | -4.02 (-13.90, 5.87) |
| **Log Insulin, pmol/L** | -0.54* (-0.81, -0.28) | 0.21 (-0.06, 0.47) | 0.12 (-0.15, 0.39) | -0.07 (-0.38, 0.25) | -0.19 (-0.51, 0.13) | 0.75* (0.49, 1.01) | -0.09 (-0.36, 0.19) |
| **HOMA-IR‡** | -0.95* (-1.36, -0.54) | 0.48* (0.07, 0.89) | 0.24 (-0.18, 0.67) | 0.07 (-0.42, 0.56) | -0.17 (-0.67, 0.33) | 1.43* (1.02, 1.84) | -0.23 (-0.66, 0.19) |
| **Ln [HOMA-IR]** | -0.75* (-1.04, -0.46) | 0.24 (-0.05, 0.53) | 0.10 (-0.19, 0.40) | -0.11 (-0.45, 0.23) | -0.21 (-0.56, 0.13) | 0.99* (0.70, 1.28) | -0.14 (-0.43, 0.16) |
| **GGT, nmol/(s*L)‡** | -22.86 (-51.31, 5.58) | -42.34* (-70.79, -13.90) | -7.21 (-36.71, 22.30) | -42.28* (-76.44, -8.12) | -35.07* (-69.70, -0.45) | -19.48 (-47.93, 8.96) | 35.14* (5.63, 64.64) |
| **Ln[GGT, nmol/(s*L)]** | -0.08 (-0.18, 0.02) | -0.11* (-0.21, -0.01) | -0.01 (-0.12, 0.09) | -0.13* (-0.25, -0.01) | -0.12 (-0.24, 0.01) | -0.03 (-0.13, 0.07) | 0.10 (-0.01, 0.20) |
| **FLI** | -15.54* (-20.62, -10.45) | -9.30* (-14.39, -4.21) | -12.22* (-17.57, -6.87) | -19.28* (-25.41, -13.18) | -7.06* (-13.31, -0.85) | 6.23* (1.14, 11.32) | -2.92 (-8.27, 2.43) |
| **hsCRP, mg/L‡** | 1.19* (0.69, 1.69) | -0.31 (-0.81, 0.19) | -0.53* (-1.04, -0.01) | -0.16 (-0.76, 0.43) | 0.36 (-0.24, 0.97) | -1.50* (-2.00, -1.00) | -0.22 (-0.73, 0.30) |
| **Ln[hsCRP, mg/L}** | 0.48* (0.13, 0.83) | -0.14 (-0.49, 0.21) | -0.39* (-0.76, -0.03) | 0.03 (-0.40, 0.45) | 0.42 (-0.01, 0.84) | -0.62* (-0.97, -0.27) | -0.25 (-0.62, 0.11) |

CI, confidence interval; BL, baseline; EOF, end-of-fast; EOR, end-of-refeed; 6wkFU, six-week follow-up; 12mFU, 12-month follow-up; BW, body weight; BMI, body mass index; kg/m2, kilogram per square meter; AC, abdominal circumference; cm, centimeter; SBP, systolic blood pressure; DBP, diastolic blood pressure; mmHg, millimeter mercury; kg, kilogram; LDL, low-density lipoprotein; HDL, high-density lipoprotein; VLDL, very-low density lipoprotein; mmol/L, millimole per liter; pmol/L, picomole per liter; HOMA-IR, homeostatic model assessment for insulin resistance; GGT, gamma-glutamyl transferase; nmol/(s·L), nanomole per second and liter; FLI; fatty liver index; hsCRP, high-sensitivity C-reactive protein; mg/L, milligram per liter; ln, natural logarithm. *Zero lies outside the 95% CI so the finding is considered significant. ‡Used robust mixed effects model on complete cases.

#### ST14. Weight Class by Visit

| Category BMI ($kg/m^2$) | BL | EOF | EOR | 6wkFU | 12mFU |
|---|---|---|---|---|---|
| | N* (%) | | | | |
| **Underweight** < 18.5 | 0 (0) | 1 (3) | 1 (3) | 0 (0) | 0(0) |
| **Normal** 18.5-24.9 | 4 (14) | 8 (28) | 8 (28) | 8 (32) | 5 (29) |
| **Overweight** 25-29.9 | 10 (34) | 11 (38) | 9 (31) | 10 (40) | 7 (41) |
| **Obese** > 30 | 15 (52) | 9 (31) | 11 (38) | 7 (28) | 5 (29) |
| **Obese I** 30-34.9 | 10 (34) | 7 (24) | 9 (31) | 7 (28) | 3 (18) |
| **Obese II** > 34.9 | 5 (17) | 2 (7) | 2 (7) | 0 (0) | 2(12) |

BMI ($kg/m^2$) reference ranges for weight categories are listed below respective variable.[4] N (%), number (percent) of participants in respective category; BMI, body mass index; $kg/m^2$, kilogram per square meter; BL, baseline; EOF, end-of-fast; EOR, end-of-refeed; 6wkFU, six-week follow-up; 12mFU, 12-month follow-up. *At the BL, EOF, EOR visits, there were 29 participants and at the 6wkFU and 12mFU visits, there were 26 and 17 participants, respectively.

#### ST15. Vital Signs by Visit

**Median (IQR)**

| | BL | EOF | EOR |
|---|---|---|---|
| **BT, °C** | | | |
| 36.1-37.2 | 36.7 (36.5, 36.9) | 36.6 (36.5, 36.7) | 36.7 (36.6, 36.9) |
| **Pulse, $min^{-1}$** | | | |
| 60-100 | 66 (60, 70) | 73 (68, 83) | 69 (64, 80) |
| **$SpO_2$, %** | | | |
| 95-100 | 98 (97, 99) | 98 (97, 99) | 98 (98, 98) |

Normal reference ranges are listed below the respective variable. IQR, interquartile range; BL, baseline; EOF, end-of-fast; EOR, end-of-refeed; 6wkFU, six-week follow up; 12mFU, 12-month follow-up. BT, body temperature; °C, degrees Celsius; $min^{-1}$, per minute; $SpO_2$, saturation of peripheral oxygen; %, percent.

#### ST16. Significance of Differences for Vital Signs

| | EOF – BL | EOR – BL | EOR – EOF |
|---|---|---|---|
| | | Estimate (95% CI) | |
| **BT, °C** | -0.06 (-0.18, 0.06) | 0.03 (-0.08, 0.15) | 0.09 (-0.02, 0.21) |
| **Pulse, $min^{-1}$** | 7.66* (3.10, 12.21) | 5.00* (0.45, 9.55) | -2.66 (-7.21, 1.90) |
| **‡$SpO_2$, %** | 0.07 (-0.56, 0.71) | 0.27 (-0.36, 0.89) | 0.19 (-0.43, 0.82) |

EOF, end-of-fast; BL, baseline; EOR, end-of-refeed; CI, confidence interval; BT, body temperature; °C, degrees Celsius; $min^{-1}$, per minute; $SpO_2$, saturation of peripheral oxygen; %, percent. *Zero lies outside the 95% CI so the finding is considered significant. ‡Used robust mixed effects model on complete cases.

#### ST17. Anti-Hypertensive Medication Use at BL, 6wkFU, and 12mFU

| | # | | |
|---|---|---|---|
| **Medications** | **BL** | **6wkFU** | **12mFU** |
| Amlodipine | 3 | 0 | 2 |
| Amlodipine and Lisinopril | 1 | 0 | 0 |
| Amlodipine and Spironolactone | 0 | 0 | 1 |
| Atenolol and Losartan | 1 | 0 | 0 |
| HCTZ† | 2 | 0 | 0† |
| Lisinopril† | 1 | 0 | - |
| Lisinopril/HTCZ*† | 2 | 0* | - |
| Losartan† | 4 | 1 | 3† |
| Propranolol | 1 | 0 | 0 |
| **Medicated Participants (% of total population)** | **15 (52)** | **1 (4)** | **6 (35)** |

At BL, 15/29 participants were taking anti-hypertensive medications, 11/15 were taking a single medication, 2/15 were taking 2 medications (Lisinopril and Amlodipine, Atenolol and Losartan) and 2/15 were taking a combination (Lisinopril/HTCZ). At 6wkFU, 1/26 was taking medication, and at 12mFU, 6/17 participants were taking medications, of which 1/6 was taking 2 medications (Amlodipine and Spironolactone). BL, baseline; 6wkFU, six-week follow-up; 12mFU, 12-month follow-up; HTCZ, Hydrochlorothiazide. *1/15 and †5/15 of the participants taking medications at BL did not participate at 6wkFU and 12mFU, respectively.

### 2. Supplementary Figures

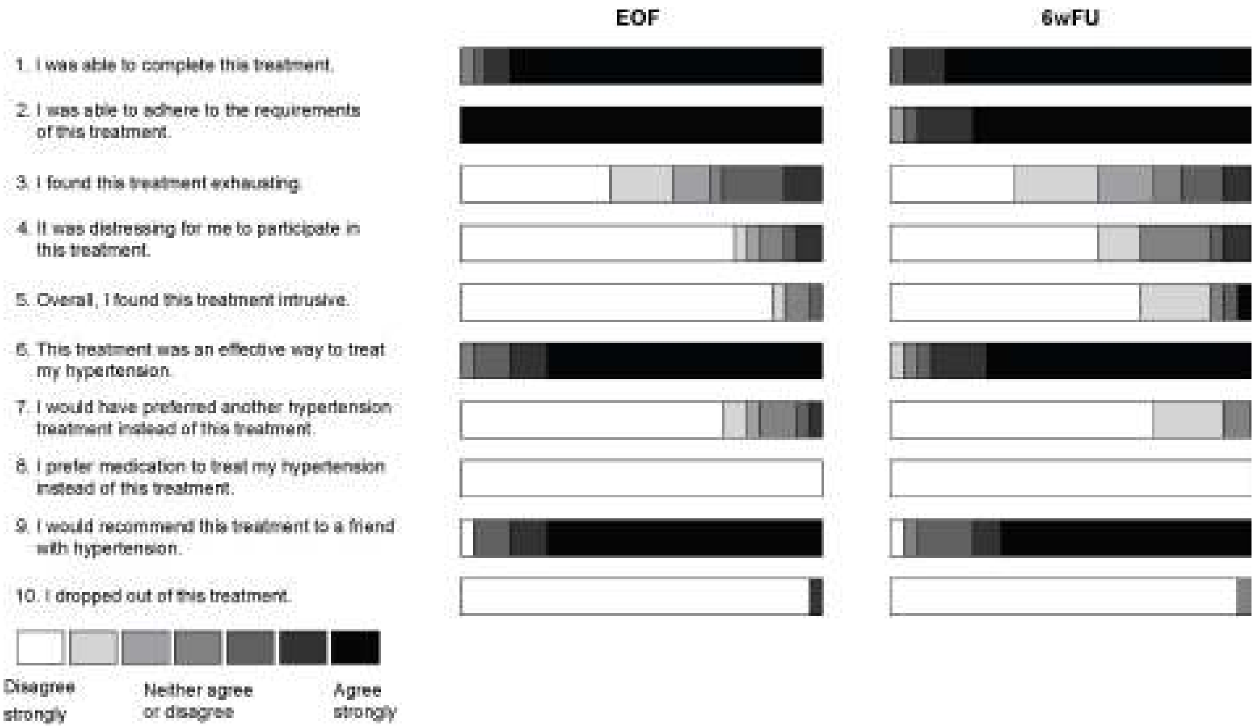


**SF1: Percentage of participants responding to individual TAAS questions**. Presented on scale from white (1, disagree strongly) to medium grey (4, neither agree nor disagree) to black (7, agree strongly). There were 29 and 26 participants at EOF and 6wkFU, respectively. EOF, end-of-fast; 6wkFU, six-week follow-up.

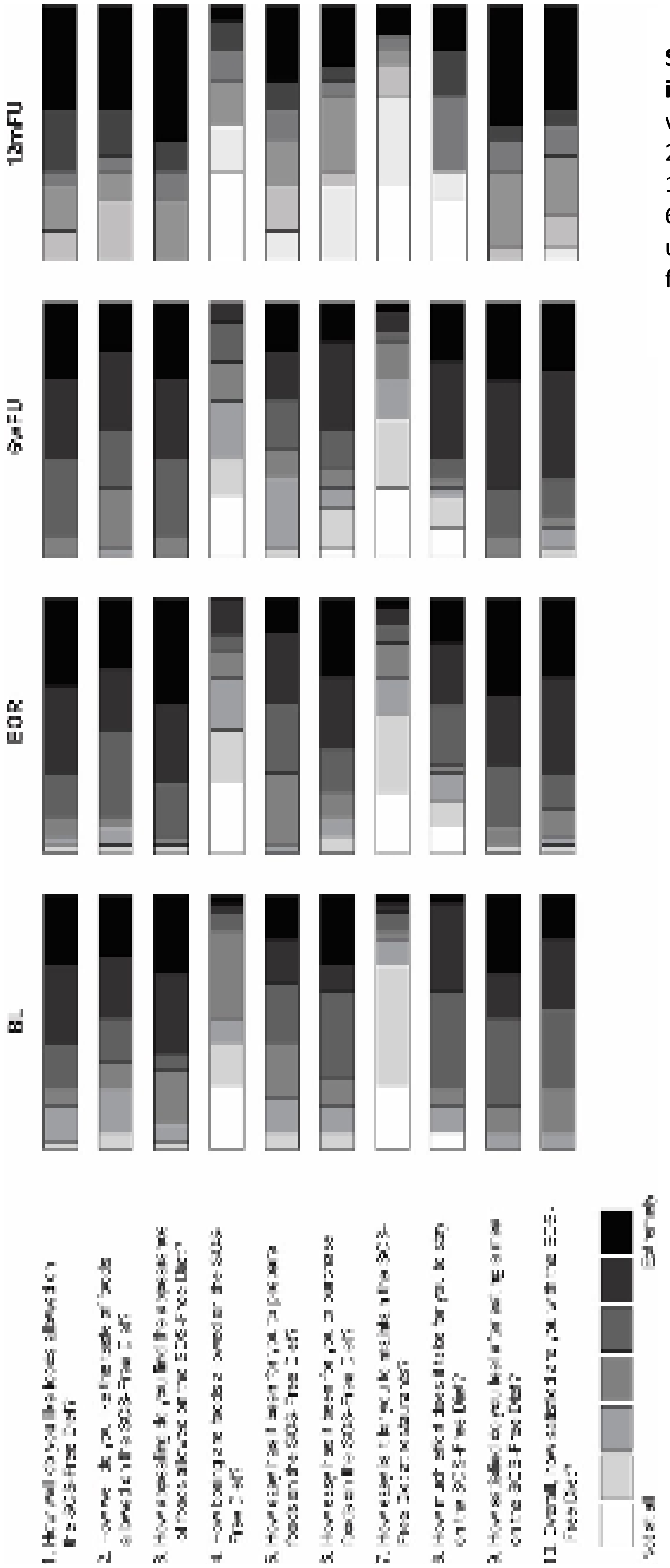


**SF2: Percentage of participants responding to individual FAQ questions**. Presented on scale from white (1, not at all) to black (7, extremely). There were 29, 29, 26, and 17 participants at BL, EOR, 6wkFU, and 12mFU, respectively. BL, baseline; EOR, end-of-refeed; 6wkFU, six-week follow up; 12mFU, 12-month follow up; SOS-Free Diet, exclusively whole-plant-food diet free of added salt, oil and sugar.

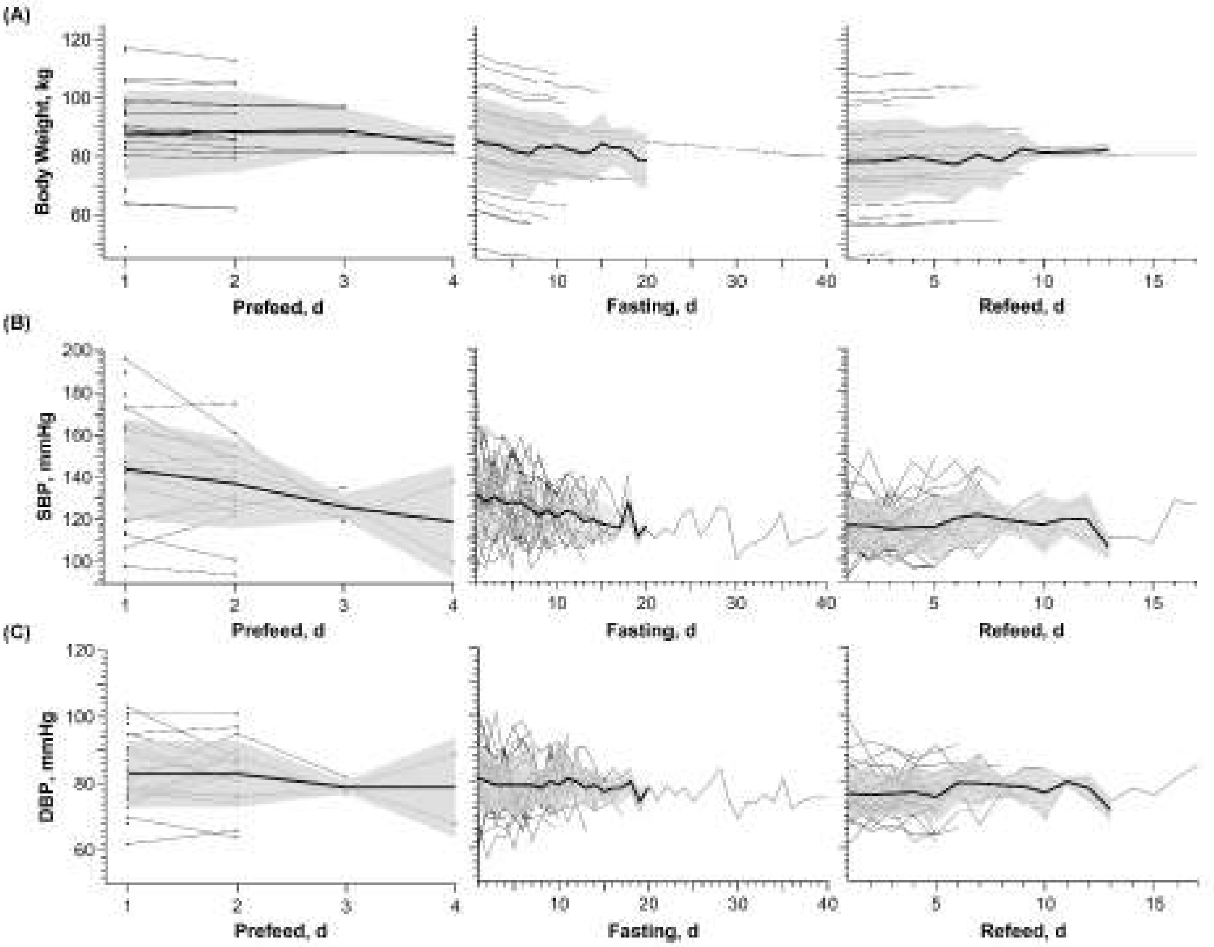


**SF3**: **Daily measurements during prefeeding, fasting, and refeeding of BW (A), SBP (B), and DBP (C)**. Values are presented as absolute values for individual participants (thin line) with overlaid mean (thick line) and stdev (grey area). SBP, systolic blood pressure, DBP, diastolic blood pressure; mmHg, millimeter mercury; d, day; kg, kilogram; stdev, standard deviation.

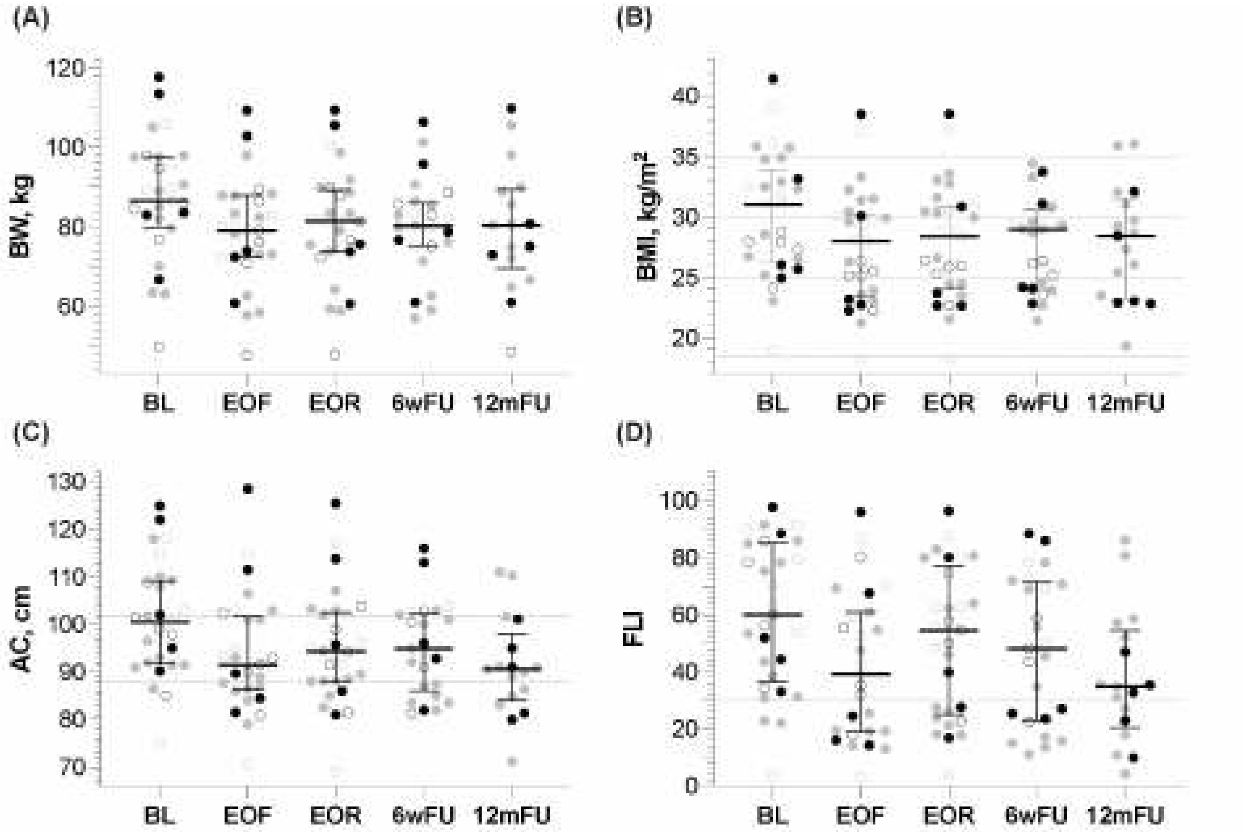


**SF4**: **BW (A), BMI (B), AC (C), and FLI (D) by visit**. Participants who attended the 12mFU visit (n=17) are depicted as full circles. Females and males are depicted as light grey and black, respectively. Horizontal dotted lines represent cut off levels for normal/optimal or abnormal values: (B) BMI categories: normal weight, 18.50 - 24.99 kg/m2; overweight, 25.00 - 29.99 kg/m2; obese, ≥ 30.00 kg/m2 (≥ 35.00, severely obese); (C) Optimal values for males ≤ 102 cm and females ≤ 88 cm; (D) FLI below 30 is optimal. BW, body weight; kg, kilogram; BMI, body mass index; kg/m2, kilogram per square meter; AC; abdominal circumference; cm, centimeter; FLI, fatty liver index; BL, baseline; EOF, end-of-fast; EOR, end-of-refeed; 6wkFU, six-week follow up; 12mFU, 12-month follow up.

#### References


[1] ICD10Data.

[2] S. Gabriel, M. Ncube, E. Zeiler, N. Thompson, M.C. Karlsen, D.M. Goldman, Z. Glavas, A. Beauchesne, E. Scharf, A.C. Goldhamer, and T.R. Myers, A Six-Week Follow-Up Study on the Sustained Effects of Prolonged Water-Only Fasting and Refeeding on Markers of Cardiometabolic Risk. Nutrients 14 (2022).

[3] LabCorp, Blood Specimens: Chemistry and Hematology.

[4] C.B. Weir, and A. Jan, BMI Classification Percentile And Cut Off Points, StatPearls, Treasure Island (FL), 2023.
